# SARS-CoV-2 waves in Europe: A 2-stratum SEIRS model solution

**DOI:** 10.1101/2020.10.09.20210146

**Authors:** Levan Djaparidze, Federico Lois

## Abstract

In order to design actionable SARS-CoV-2 strategies, we extended the SEIRS model to support stratified isolation levels for healthy <60 and vulnerable individuals. At first, we forced isolation levels to be uniform, showing that daily deaths curves of all metropolitan areas in the analysis can be fitted using homogeneous Ro=3.3. In the process, we established the possibility that an extremely short infectiousness period of 2 days coupled with 5 days exposure may be responsible for the multiple deaths valleys observed during the weeks following lockdowns. Regardless of the infectiousness period, we realized that is possible to infer non-uniform isolation levels for healthy <60 and vulnerable by forcing the model to match the <60 to >60 age serology ratio reported in seroprevalence studies. Since the serology ratio is more robust than absolute values, we argue immunity level estimations made in this way (Madrid 41%; Catalonia 23%; Brussels 49%; and Stockholm 62%) are closer to reality. In locations where we didn’t find reliable serology, we performed immunity estimations assuming Spain’s serology ratio (Paris: 23%; London: 33%). We predict that no location can return to normal life without having a second wave (albeit in Stockholm a smaller one). We searched what isolation values allow to return to normal life in 90 days minimizing final deaths, shockingly all found isolations for healthy <60 were negative (i.e. coronavirus parties minimize final deaths). Then, assuming an ideal 1-day long vaccination campaign with a 77% efficacy vaccine, we compared predicted final deaths of those 90-day strategies for all possible vaccination dates with a 180-day long vaccine waiting strategy that imposes 0.40 mandatory isolation to healthy <60 and results in 0.65 isolation to vulnerable. We found that 180-day of mandatory isolations to healthy <60 (i.e. schools and workplaces closed) produces more final deaths if the vaccination date is later than (Madrid: Feb 23 2021; Catalonia: Dec 28 2020; Brussels Apr 25 2021; Paris: Jan 14 2021; London: Jan 22 2021). We also modeled how average isolation levels change the probability of getting infected for a single individual that isolates differently than average. That led us to realize disease damages to third parties due to virus spreading can be calculated and to postulate that an individual has the right to avoid mandatory isolation during epidemics (SARS-CoV-2 or any other) if these damages can be covered with a novel proposed isolation exemption insurance policy. As secondary findings in Appendix III we hypothesize that an early D614 like strain wave might be the cause of low mortality in Asia, and show the negligible reduction of HIT due to heterogeneity. Finally we conclude that our 2-stratum SEIRS model is suitable to predict SARS-CoV-2 epidemic behavior and can be used to minimize covid-19 disease and isolations related damages. To the sole effect of understanding and verifying its content the same model used through this paper has been made available online at www.sars2seir.com/paper-12-2020/

## Background

Since the SARS-CoV-2 virus pandemic showed the potential to overwhelm health systems, harsh restrictions were imposed on individuals around the world. Early data from South Korea indicated that the infection fatality rate (IFR) is much higher for vulnerable individuals (i.e. >60 and <60 with underlying diseases) than for healthy <60. This led (5) early in March to propose a vertical quarantine and eventually a growing number of countries implemented it. But with the notable exception of Sweden, governments did not take advantage of the fact that IFR for healthy <60 is not only lower than for vulnerable individuals but, as was pointed out early on by the author of (6), is also negligible.

The computer epidemiological model of the Imperial College (2) and the IHME tool were initially used to predict the epidemic behavior but they did not model stratified isolations and suffered skepticism for their predictions (3)(4).

Sweden’s lead epidemiologist (Dr. Anders Tegnell) designed a strategy that did not impede viral spread among healthy <60. In theory (sustainability aside) the strategy could allow speedily reaching HIT without infecting as many vulnerable individuals. If successful, it would mean that closing schools and workplaces not only is economically damaging but also sub-optimal in the sense that it results in a higher death count since vulnerable individuals cannot sustainably isolate for long periods.

This strategy has been questioned because seroprevalence studies detected only 7% of the population with SARS-CoV-2 antibodies in Stockholm (7) similar to the percentages observed in other countries that implemented stricter lockdowns (e.g. average seroprevalence in Spain was 5%) (8).

Searching for hidden immune response, authors in (15) found that 35% of asymptomatic individuals with SARS-CoV-2 T cell response did not have detectable antibody titer levels. This means that antibody tests underestimate immune response seroprevalence, but even using a uniform 45% proportion of asymptomatic to correct the underestimation, the average seroprevalence value only increases 18% (i.e. from 7.0% to 8.3%, still far from HIT). Of course, diminishing titer levels (33) and test sensitivity can explain low reported serology levels, but why did daily deaths also ease in Madrid? Can it have the same immunity level as Stockholm?

Even though average serology reported in May were similar, <60 / >60 age serology ratio in Madrid (i.e. 4.78/6.03=0.79) was much lower than in Stockholm (i.e. 6.09/3.58=1.70). This means that proportionally to <60 individuals, many more >60 individuals were infected in Madrid than in Stockholm. Since the death count mostly comes from >60, a higher age serology ratio in Stockholm necessarily means higher immunity level (once death count is adjusted by the population pyramid).

In June, the Public Health Agency of Sweden (9) used a Susceptible->Exposed->Infected->Recovered (SEIR) model to estimate immunity levels (i.e. total recovered / population). Estimations and predictions for Stockholm were 13% (May 1st), 19% (July 1st), and 20% (September 1st). Daily reported positive cases curve fitting was accomplished using a sophisticated time-varying infectivity rate. But as the authors warned, the model did not have age-stratified isolation levels, and since the serology ratio in Stockholm was higher than 1 the real immunity level has to have been underestimated.

Also using an SEIR model, authors of (10) were able to successfully fit ICU hospitalizations, but also did it without modeling stratified isolations. In (11) an n-rank matrix of cross isolations between groups (i.e. instead the 2-rank used here) was proposed to demonstrate how stratified age isolations could result in lower death count but they did not test their SIR-like model with real data curves.

A potentially infectivity increasing mutation of the SARS-CoV-2 virus (i.e. G614 variant) was reported in (37), but even though it seems to be more infective than an earlier D614 variant, no increase in disease severity was reported (36). As pointed out in (38) not every mutation deserves to be classified as a strain, but it will eventually happen and thus it should be modeled.

Vanishing waves across Europe have led authors in (12) to postulate Ro heterogeneity across the population to justify a hypothetical HIT lower than the initial estimation of 70%, thus implying that normal life can return without reaching 70% immunity level. The proposal is ingenious, but evidence does not show the heterogeneity impact due to susceptibility that would most likely exist between <60 and >60 individuals. Countries with young populations (e.g. Brazil) have had higher (initially unrestricted) Ro estimations (13) than Europe, although this higher Ro could be the result of an initial lower prevalence of the less infective D614 strain in Brazil than in Europe. Nevertheless, and even though deaths curves can be fitted using a homogeneous Ro (e.g. 3.3 implying HIT 70%) and two non-uniform isolation levels (that is: using only effective temporal R heterogeneous connectivity in the context of (12)), we modeled heterogeneity due to non-contagious asymptomatic and lower connectivity, results are shown in Appendix 3: Negligible reduction of HIT due to heterogeneity.

Early cases from South Korea pointed to short-lived immunity and non-negligible re-infection rate, but after revision, those cases were discarded as re-infections (34). The same report discarded the possibility of the virus entering cell nucleolus and thus becoming chronic via that mechanism. Eventually new findings may point to non-negligible probability of re-infection and hence it is worth being modeled (i.e. SEIRS).

With all that in mind, we extended the classic SEIRS model to support stratified isolation but using only two isolation levels, one for healthy <60 and another for vulnerable (2-stratum). We had the intention to fit daily deaths curves maintaining virus parameters (e.g. IFR, Ro, exposure, infectiousness, etc.) constant through all locations and also maintaining isolation values constant during months within each location. We think that to have credible predictive value, constant for months isolation levels should be able to average every other second-order influence in the epidemic behavior.

We developed an algorithm that allowed us to easily fit reported daily deaths curves (total, peak, and shape) by automatically adjusting isolation to vulnerable and the implicit amount of infected in a hypothetical first non-communitarian infection date. A single attempt to fit reported deaths without this tool is enough to see how handy it is.

Originally we were set to treat isolation to healthy <60 as a plausible value (e.g. “in Stockholm was lower because schools were open”) but rapid fitting enabled us to realize (eventually) that it was possible to disambiguate non-uniform isolations by matching the age serology ratio predicted by the model with the age serology ratio reported in seroprevalence studies.

Almost every other finding in this paper also came from this tool: Immune level estimation sensitivity analysis (e.g. change Ro and fit); Estimating the spreading day if only one non-communitarian spreader is assumed (i.e. change initial spreading day until fitted initial spreaders is 1); Finding that Do=2 coupled with Eo=5 can explain the multiple valleys after lockdowns observed in the 1-day moving average daily death curves (i.e. fitting a dozen of different pairs of Do and Eo); Include lack of IgG in asymptomatic when predicting reported serology ratio (i.e. add TAK variable and equation); Estimate the proportion of asymptomatic for <60 and >60 (i.e. change proportions and fit until predicted asymptomatic to symptomatic ratio in Spain matches the reported = 0.51); Estimate that the proportion of SARS-CoV-2 positive reported deaths that are “with” the virus is 15% (i.e. add testing positive period parameter and yearly probability of dying for other causes to the predicted sars-cov2 positive deaths and lower IFR_vul until predicted serology ratio matches again); Estimate the minimum value of IFR_vul to have another wave in Brussels or the maximum that avoids the low one in Stockholm (i.e. change IFR_vul and fit until the second wave appears or disappears); Hypothesize that an early wave of a D614 like variant can be the cause of low mortality in most locations in Asia (i.e. fit with an hypothetical lower IFR_vul competing strain and then simulate setting the dominant spreading event 60 days earlier); Estimate HIT reduction due to heterogeneity (i.e. under heterogeneity change Ro until I_dayS value is the same that under homogeneity).

We also developed a death minimizing algorithm with the original intention of testing the general intuition that not impeding spreading among healthy <60 not only allows returning to normal life earlier but also can result in lower final deaths. Yet again, the tool led us to unexpected findings: Initial “Back in March 2020” death minimizing strategies included low isolation to healthy <60 (and also would have resulted in manageable ICU occupancy); After sub-optimal fitted strategies isolation to healthy <60 death minimizing values are negative (it took us a couple of weeks to let the algorithm search for values lower than 0.00); When a vaccine with sufficient efficacy is available in the future given a long enough strategy and a high enough isolation to vulnerable the death minimizing value for healthy <60 is the maximum available (i.e. the strategy pushes the wave to after the vaccine). That last result was not unexpected but it was revealing to see how the algorithm changed of strategy if the vaccination date has just one day of difference. That led us to compare 90-day death minimizing strategies against a tentative 180-day long vaccine waiting strategy for all possible vaccination dates and found the risk of final deaths increase if vaccination does not occur before a specific date.

The 2-stratum SEIRS model is an iteration of simple algebraic operations, but its behavior is highly non-linear and the automatic fitting and death minimizing algorithms proved to be very helpful for modeling real locations.

## 2-stratum SEIRS model

Table 1 details all virus common parameters. See Appendix II for point estimates discussion and Table A.1 for immune level estimation sensitivity.

**Table 1.** Virus common parameters description and values

| Model Variable | Value | Description |
| --- | --- | --- |
| $R_0$ | 3.3 | Reproduction number |
| $E_0$ | 5 | Exposure total duration (days) |
| $D_0$ | 2 | Infectiousness average duration (days) |
| $R_{toD}$ | 11 | Recovered to death period (days) |
| $S_0$ | 1.00 | Proportion of susceptible population |
| IFR_vul | 0.0093 | Infection fatality rate for vulnerable |
| IFR_non_vul | 0.00048 | Infection fatality rate for healthy <60 |
| $P_{vul\_u60}$ | 0.0342 | % of <60 that are vulnerable |
| $P_{non\_vul\_una}$ | 0.07 | % of healthy <60 unable to isolate from vulnerable |
| $T_1$ | 180 | Period between recovered and losing immunity (days) |
| PRLI | 0.00 | Proportion of recovered losing immunity after $T_1$ |
| $A_{o60}$ | 0.21 | % of >60 that are asymptomatic |
| $A_{u60}$ | 0.52 | % of <60 that are asymptomatic |
| TAK | 0.35 | % of asymptomatic with T cells and without IgG |
| APTP | 17 | Average positive testing period (days) |
| $Ma$ | 7 | Daily deaths moving average (days) |
| ICU_pd | 0.45 | Average ICU hospitalizations per death |
| ICU_h_dur | 10 | Average ICU hospitalization duration in (days) |
| vEff | 0.77 | Vaccine efficacy (uniform) |

As depicted in (14) isolation strategies are not purely age-stratified. In the vulnerable isolation group we include >60 population (i.e. P_o60 parameter) and also <60 with underlying diseases (i.e. P_vul_u60). And since a proportion of healthy <60 may not be able to isolate from vulnerable (i.e. P_non_vul_una) they are also isolated in the same group. Inside the model’s code they are called the “unable” (i.e. unable to isolate from vulnerable, see Appendix I – SEIRS model with 2-stratum isolation), but through this paper we call them the vulnerable for short. Modeling more than 1-stratum for <60 may only add second-order predicting value because of the high proportion of young age people living with their <60 parents in the same household (i.e. too high P_non_vul_una for the additional stratum).

And to estimate deaths, IFR_vul cannot simply be multiplied by the recovered from the “unable” compartment because healthy <60 isolating with them have different infection fatality rate (see nD_vul formula in Appendix I).

### Daily deaths fitting

We were able to fit all locations to match curve shape, peak (7-day moving average), and total sars-cov2 positive daily reported deaths using uniform isolation parameters (See Table 5). Virus parameters were common to all locations and also constant during the isolation phase (see Table 1). Location-specific data is well known and was also constant during the simulation (see Tables 2 and 3). See Appendix II for the location-specific parameters discussion and Table A.2 for immune level estimation sensitivity analysis.

**Table 2.** Location-specific variables description

| Model Variable | Description |
| --- | --- |
| Pop | Population |
| P_o60 | % of population >60 |
| Iso_1_real | Initial day of lockdown (from to 12/01/19) |
| con_oD | Reported daily SARS-CoV-2 deaths |
| sero_u60_o60 | <60 to >60 reported serology ratio |
| sero_day | Day of serology study (from 12/01/19) |
| PYDR_vul | Population yearly death rate for vulnerable individuals |
| PYDR_non_vul | Population yearly death rate for non-vulnerable individuals |

**Table 3.** Location-specific values

| Location | Stock-holm | Madrid | Catalo-nia | Brussels | Paris | London |
| --- | --- | --- | --- | --- | --- | --- |
| Population | 2.38M | 6.66M | 7.73M | 1.21M | 12.1M | 10.6M |
| Pop >60 (%) | 21.0% | 23.2% | 24.8% | 17.6% | 19.8% | 20.0% |
| Lockdown day | 3/24/20 | 3/09/20 | 3/12/20 | 3/19/20 | 3/17/20 | 3/16/20 |
| Daily deaths (per million at end of 1st phase) | SARS-CoV-2 positive reported daily deaths (date of deceased) |  |  |  |  |  |
|  | (968) | (1259) | (734) | (854) | (620) | (760) |
| Serology day | 5/03/20 | 7/05/20 | 7/11/20 | 4/26/20 | NA | NA |
| Reported serology ratio <60/>60 | 1.70<br>6.16/3.64 | 0.79<br>4.78/6.03 | 0.79<br>4.78/6.05 | 1.21<br>6.13/5.06 | NA | NA |

**Table 4.** Fitting specific parameters description

| Model Variable | Description |
| --- | --- |
| S_day | Day of the first infection (from 12/01/19) |
| I_dayS | Infected in S_day |
| Iso_1_dur | 1st phase isolation duration (days). |
| Iso_1_res_abl | 1st phase isolation level for healthy <60 |
| Iso_1_res_una | 1st phase isolation level for vulnerable |

**Table 5.** 1st phase uniform isolation biased immunity level estimations

| Location | Stock-holm | Madrid | Catalo-nia | Brussels | Paris | London |
| --- | --- | --- | --- | --- | --- | --- |
| Uniform isolations values | 0.675 | 0.680 | 0.728 | 0.695 | 0.72 | 0.71 |
| Predicted serology ratio and (reported) | 0.88<br>(1.70) | 0.88<br>(0.79) | 0.88<br>(0.79) | 0.88<br>(1.21) | 0.88 | 0.88 |
| Biased immunity estimation<br>R / pop | 38% | 45% | 25% | 38% | 25% | 36% |

If isolation levels are forced to be equal (i.e. Iso_1_res_abl = Iso_1_res_una), the daily deaths curve can be fitted but predicted serology ratios do not match reported serology ratios and hence immunity estimations are biased (Table 5):

This demonstrates that disregarding serology ratio, waves across Europe can be fairly fitted with homogeneous Ro = 3.3 and using only one uniform isolation (ranging from 0.675 in Stockholm to 0.728 in Catalonia) constant for several months (see 1st isolation duration values in Table 6).

**Table 6.** Immunity levels estimated with 1-phase stratified isolation levels matching predicted and reported <60 / >60 serology ratio.

| Metropolitan area | Stockholm | Madrid | Catalonia | Brussels | Paris* | London* |
| --- | --- | --- | --- | --- | --- | --- |
| Initial spreading day | 1/11/20 | 12/19/19 | 12/25/19 | 1/9/20 | 12/28/19 | 12/25/19 |
| Infected on initial spreading | 1.03 | 1.10 | 1.04 | 1.14 | 1.07 | 1.00 |
| 1st isolation duration (days) | 95 | 102 | 100 | 101 | 113 | 104 |
| Isolation to Vulnerable | 0.941 | 0.588 | 0.654 | 0.872 | 0.641 | 0.619 |
| Isolation to healthy <60 | 0.3 | 0.748 | 0.78 | 0.53 | 0.765 | 0.76 |
| Predicted serology ratio | 1.70 | 0.79 | 0.79 | 1.21 | 0.79 | 0.79 |
| Immunity estimation date | 6/30/20 | 7/06/20 | 7/07/20 | 7/15/20 | 7/25/20 | 7/15/20 |
| Immunity level estimation R / pop | 62% | 41% | 23% | 49% | 23%<br>(with Spain's serology) | 33%<br>(with Spain's serology) |

### Immunity level estimation

Forcing the simulation to match the reported serology ratio disambiguates the isolation levels for vulnerable and healthy <60 and changes immunity level estimation (Table 6), but still allows to fit reported daily deaths (see reported and predicted curves coinciding in Graphics 1.1 to 1.6). We maintain the immunity levels found in this way are closer to reality.

Now we can answer the initial question, can Madrid have the same immunity level as Stockholm? The answer is no. Stockholm is closer to HIT than Madrid. And we can also predict that no location can return to normal life without having a second wave (Graphics 1.1 to 1.6).

We did not find reliable serology ratios prior to July in London and Paris. Instead of using uniform isolations, we approximated for those locations with the serology ratio reported for Spain (i.e. 0.79). In tables or graphics where this assumption was deployed we mark the location with an asterisk (*).

Are there other field ratio values that could be used to disambiguate isolation values? <60 to >60 confirmed case ratio (e.g. PCR) cannot be matched to <60 to >60 infections ratio produced by the model because <60 are sub sampled at hospital gates. And <60 to >60 deaths ratio cannot be used either because healthy <60 deaths are negligible and the model assumes that <60 vulnerable individuals are isolated together with >60, so isolation changes barely affect the <60 to >60 deaths ratio predicted by the model. How is possible that in Madrid isolation to vulnerable was lower than isolation to healthy <60? Even after correcting for lack of IgG in asymptomatic (see Appendix III - Secondary findings, Asymptomatic: 52% of <60 and 21% of >60) age serology ratio was 0.90, meaning vulnerable were infected in a higher proportion that healthy <60. One explanation could be that individuals inside nursing homes were not able to isolate properly and, even though they are a small proportion of the vulnerable population (796 beds / 100.000 inhabitants), high infection rates were enough to produce a seroprevalence ratio < 1. Isolations could be fitting just that.

See Appendix II for the detailed analysis of the immunity level sensitivity to the model’s parameters. Summarizing, other than the obvious sensitivity of So: -2.44 (see effects of immunity build prior to March 2019 in Appendix III - Early D614 like strain wave in Asia: A hypothesis), the only common virus parameter with relevant sensitivity is IFR_vul: -0.85. Of the population parameters also sero_ratio: +0.75 has relevant sensitivity, but surprisingly both of them have lower sensitivity than trivial population parameters like pop: -1.23, pop_o60: -1.00, and con_oD: +0.98. Immune level estimations are more sensitive to the exact amount of population from where reported deaths are drawn than to IFR_vul or seroprevalence ratio.

### Predicting second waves

What locations can return to normal life? Fitted + normal life was simulated with fitted 1st-phase isolations values and thereafter all isolations set to 0. Graphics 1.1, 1.2, 1.3, 1.4, and 1.6 show how nearly uniform isolation levels managed to extinguish the death curves but resulted in an immunity level much lower than HIT and hence Madrid, Catalonia, Brussels, Paris*, and London* are set for another wave.

**Graphic 1.1.**
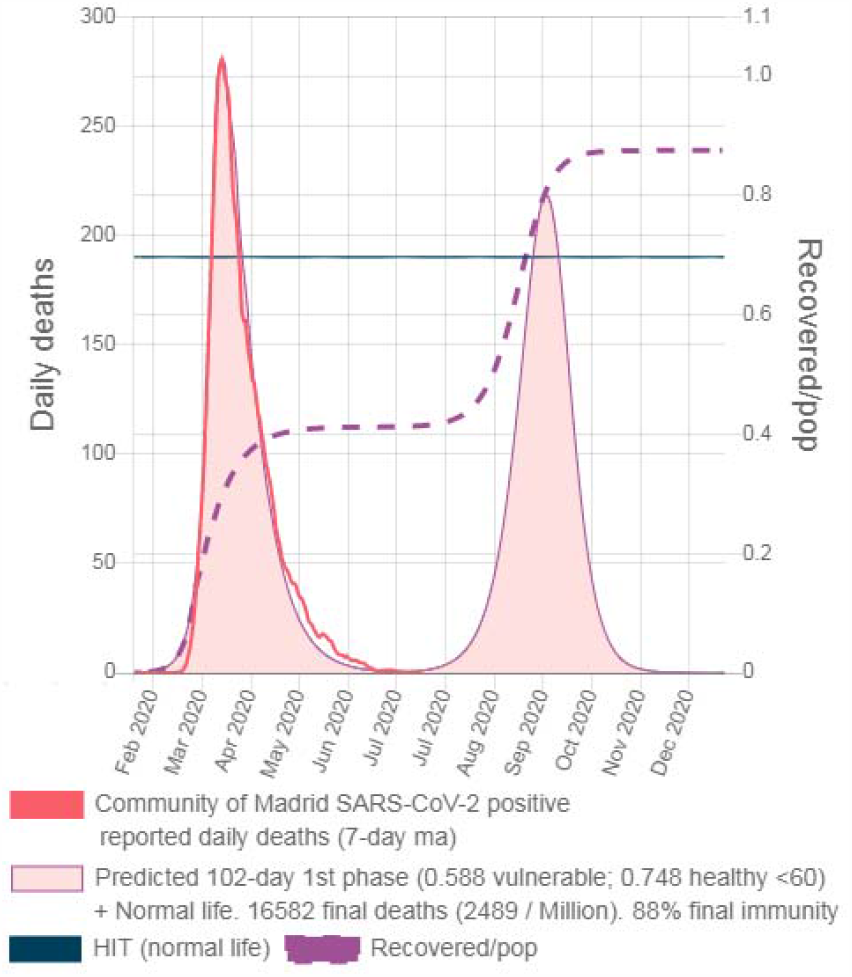
Madrid fitted isolations + Normal Life

**Graphic 1.2.**
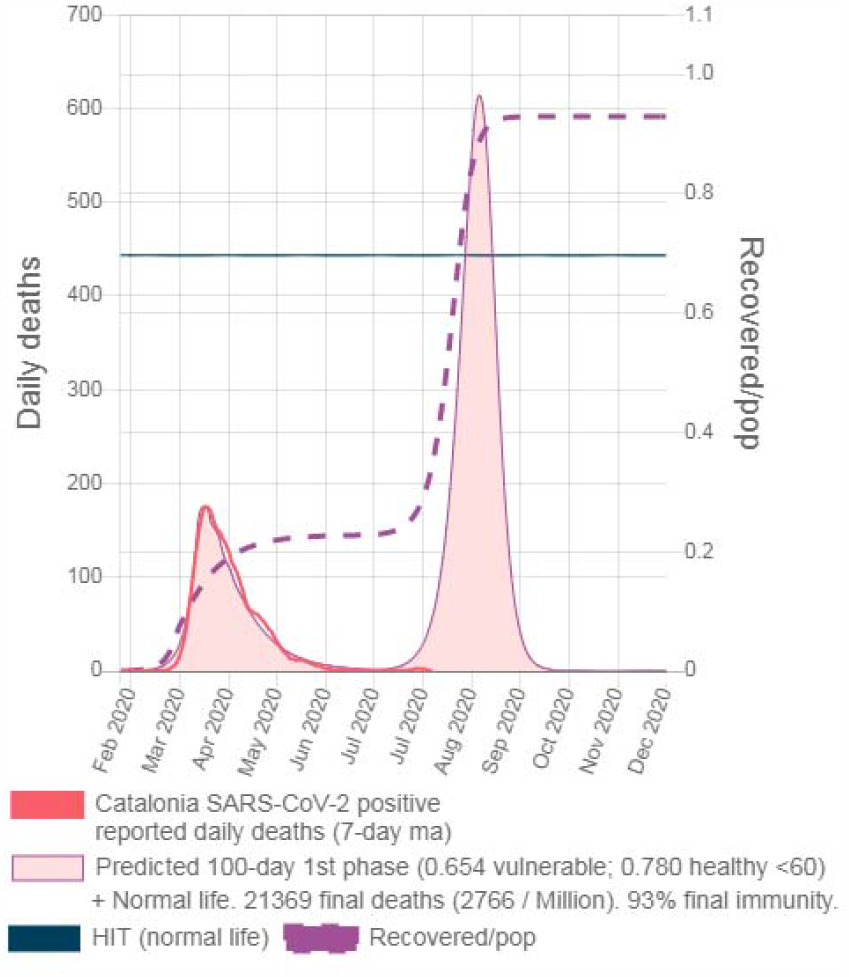
Catalonia fitted + Normal Life

**Graphic 1.3.**
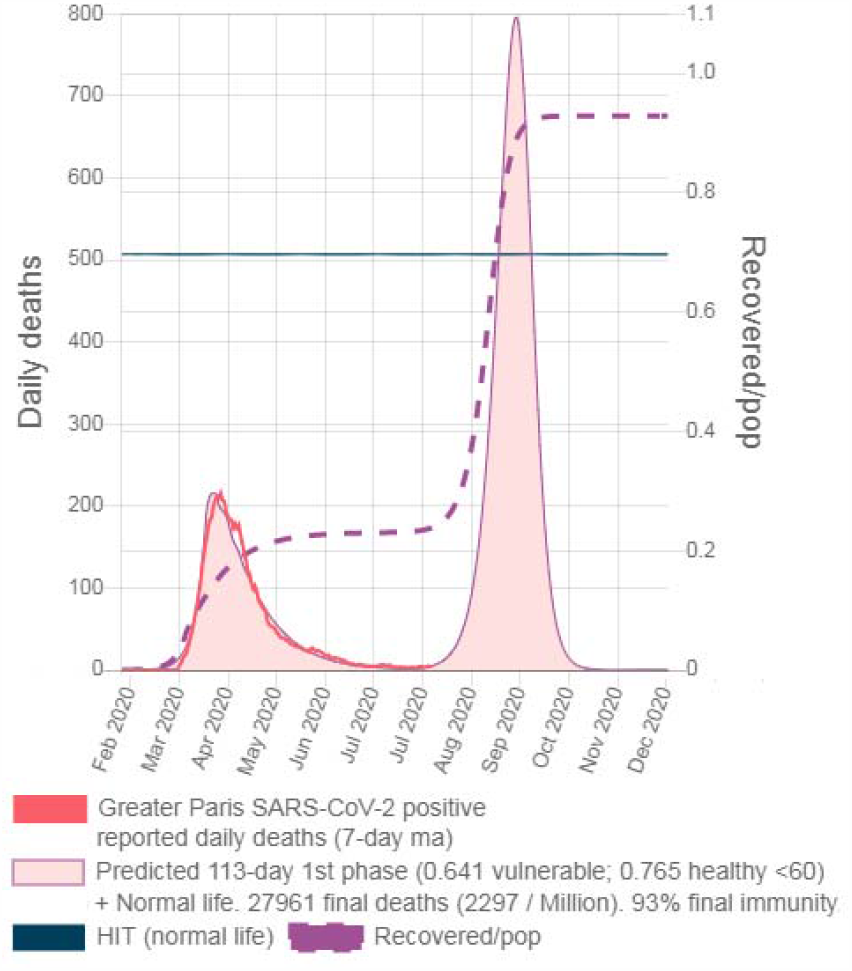
Paris* fitted + Normal Life

**Graphic 1.4.**
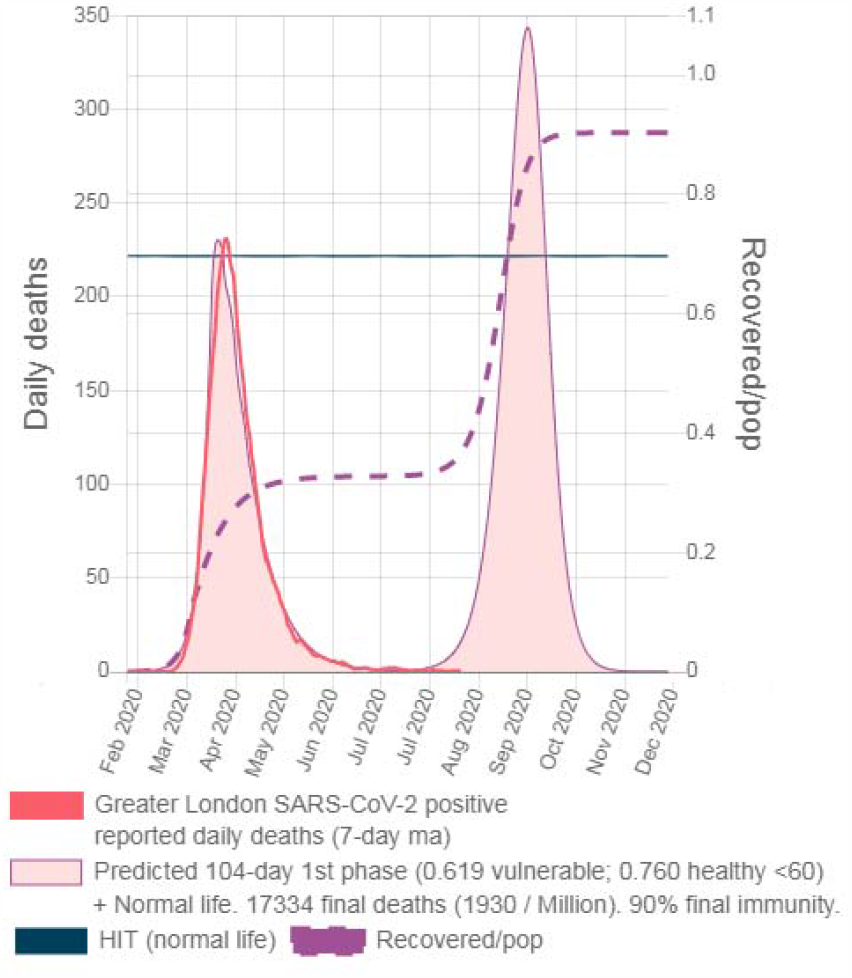
London* fitted + Normal Life

**Graphic 1.5.**
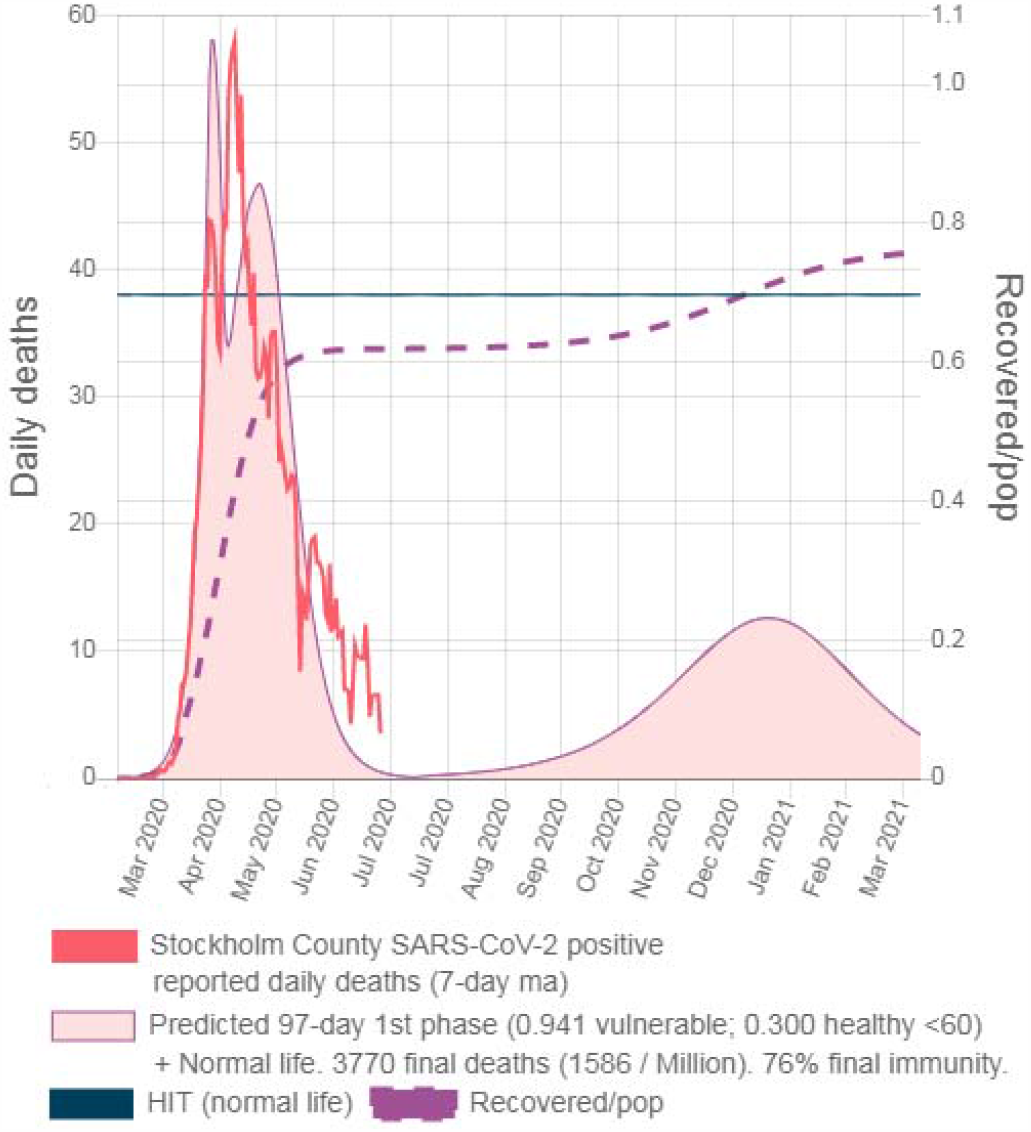
Stockholm fitted + Normal Life (smaller second wave peaking in Jan 2021)

**Graphic 1.6.**
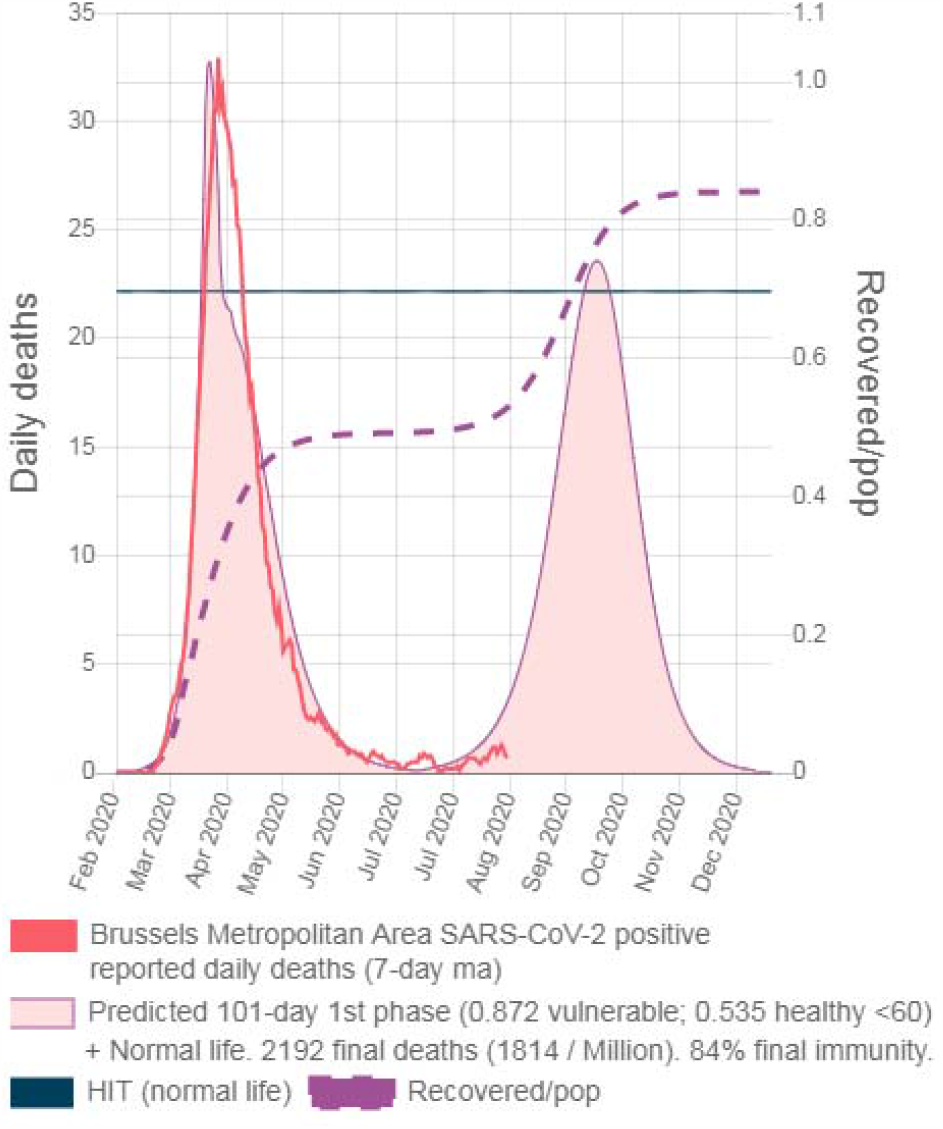
Brussels fitted + Normal Life

Stockholm (Graphic 1.5) is close to HIT but still has second wave (smaller than the first one) peaking on Jan 2021. The simulation introduces 1 infected every 10 days after 60 days from lockdown (to mimic some minimal additional non-communitarian disturbances) but if these disturbances are higher in real life the second wave would occur sooner. Stockholm won’t have a second wave if IFR_vul < 0.81%.

Since in Brussels only 69% of daily reported deaths are SARS-CoV-2 positive deaths (43), then parameter con_oD_scale=0.69, fitted deaths per million decreases from prior value of 1238 to 854, immunity level estimation decreases from 70% to 49% and therefore Brussels is also set for another wave (see Graphic 1.6). Reported serology ratio is 1.21 and the curve shape still remains compatible with a lower value.

Since we have inferred exposed on initial spreading day (i.e. most of them are in late December 2019 if 1 single non-communitarian spreader is assumed) we can use it to predict what would have been the outcomes of applying other initial strategies. “Do nothing” strategy is simulated with all isolations set to 0.00 and “The Sweden Strategy” is simulated with 90-day isolations of 0.941 to vulnerable and 0.30 to healthy <60 (initiating on lockdown day).

In Graphic 2.1 it can be seen how fast-spreading due to no isolations in London would have overshot immunity level to 97% well above theoric HIT (i.e. 70%). In Graphic 2.2 it can be seen how even with a 0.94 value of isolation to vulnerable overshooting among healthy <60 would have allowed almost reaching HIT.

ICU demand and overall deaths predicted by the Imperial College Model in (2) were initially too high because the average IFR used was too high (i.e. 0.84% average for all ages) and implicit values for London used here are closer to 0.15% (i.e. 0.92% for vulnerable and 0.0035% for non-vulnerable with P_o60 = 0.1643), but “Do Nothing” strategies would have overwhelmed unprepared health systems anyway (+45 ICU/100K in every location).

**Graphic 2.1.**
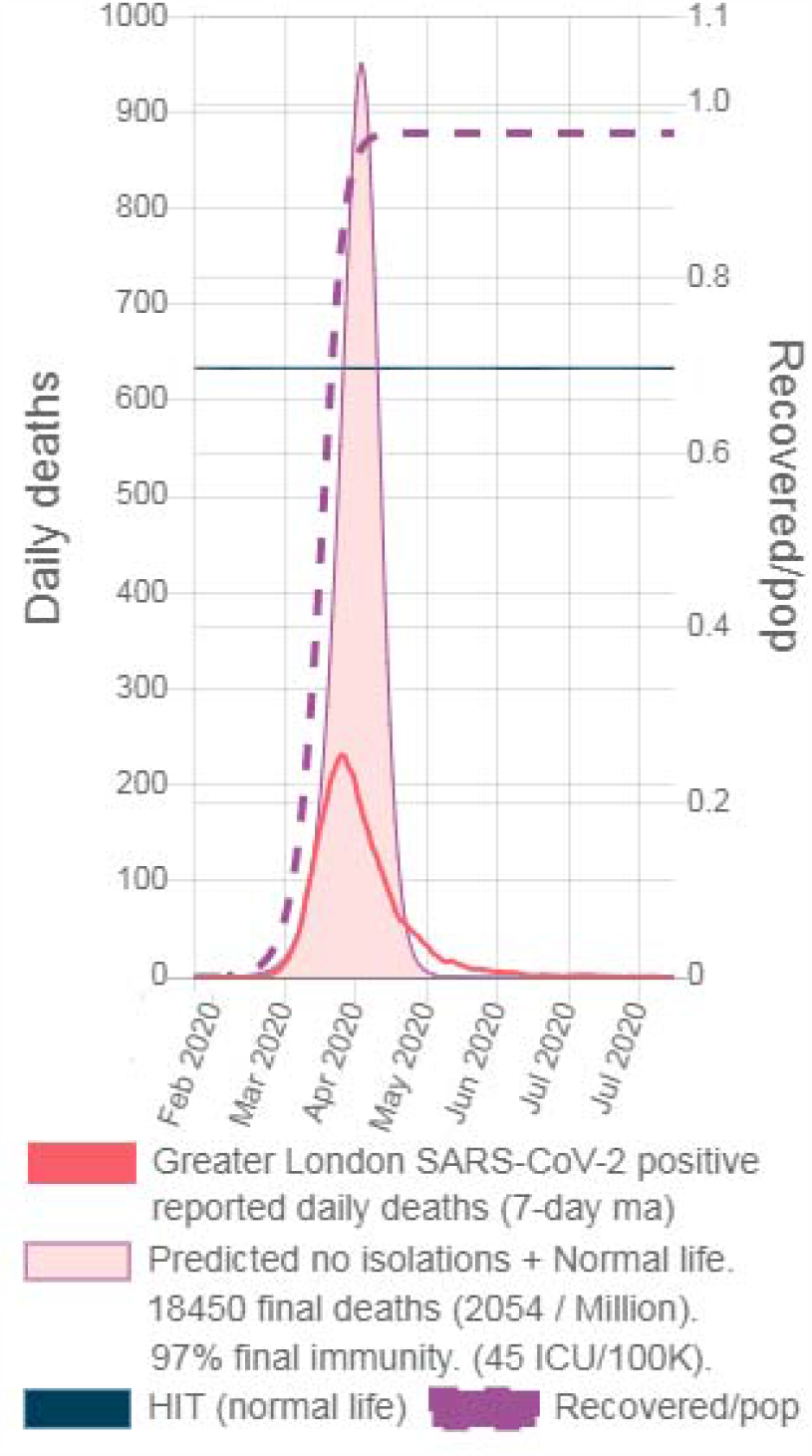
“Do nothing” + Normal Life in Greater London

**Graphic 2.2.**
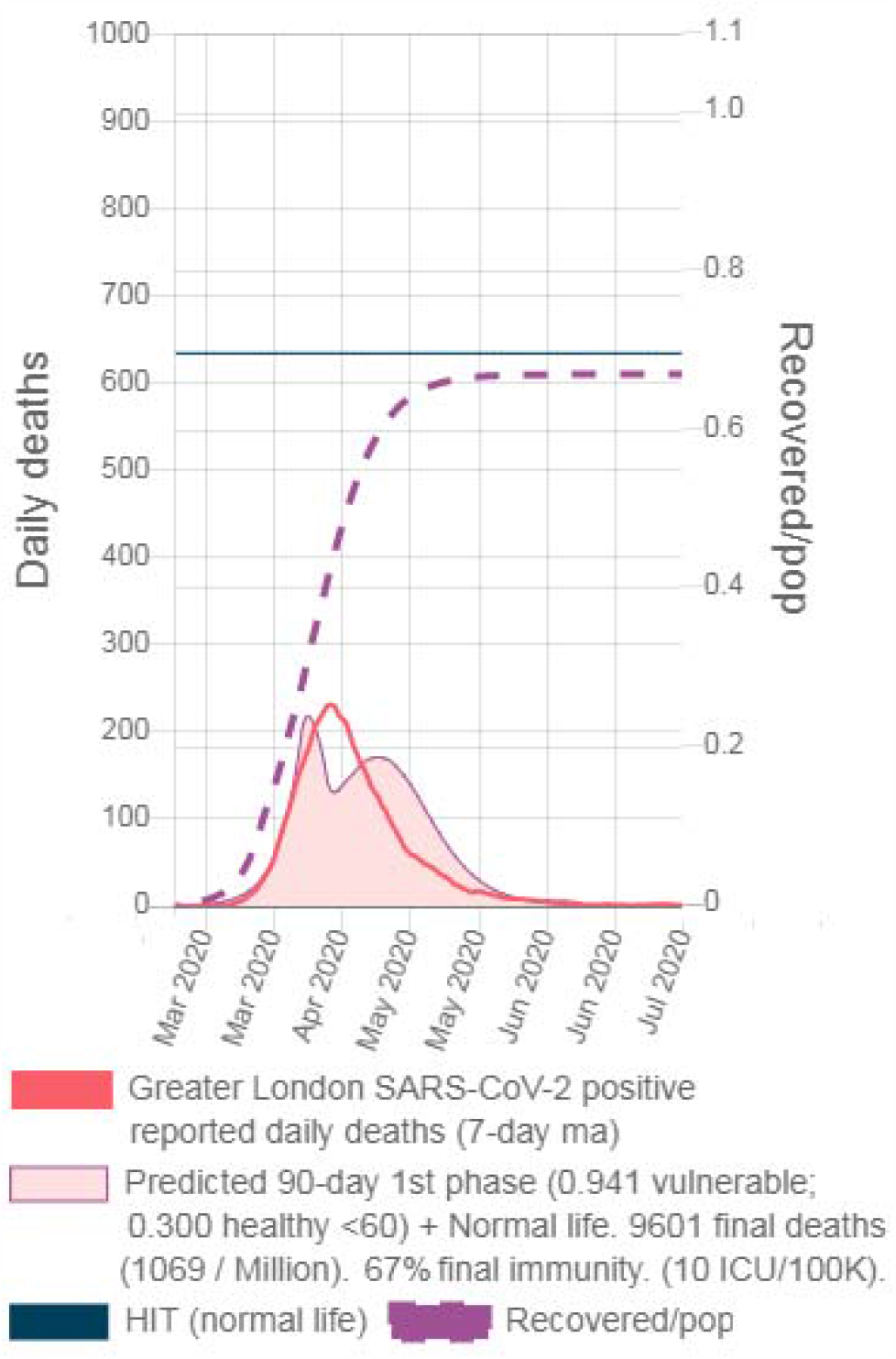
Sweden’s strategy + Normal Life in Greater London

Table 7 demonstrates that a 90-day initial strategy with 0.941 and 0.30 isolation values (i.e. Sweden’s fitted strategy) would have allowed returning to normal life with less final deaths than the predicted for the fitted nearly uniform isolations (and also with less ICU utilization). How close to optimum was Sweden’s strategy?

**Table 7.**
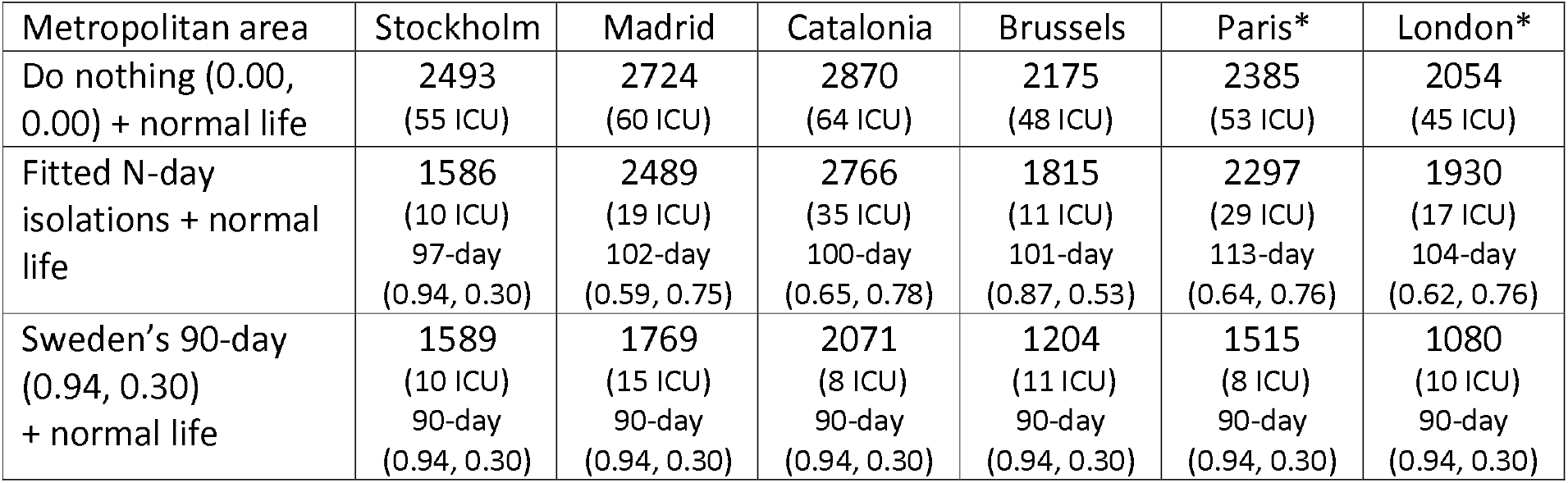
Final deaths per million and ICU/100K utilization for various past strategies (beginning on lockdown day) and followed by normal life

### Deaths minimizing strategies

No location can return to normal life without having another wave and governments are set for another round of mandatory policies. What are the alternatives?

The Test/Trace/Isolate (TETRIS) strategy has been proposed to stop the virus. But since pre-symptomatic are found to be infectious (25) tracing back contacts in a pure TETRIS scenario without mandatory isolation is impossible (i.e. imagine contagions in public night clubs). And, since mild symptomatic are also infectious (26) and they do not usually report the disease, even when applied together with harsh isolations TETRIS becomes impracticable after a couple hundred of contagions (i.e. virus spreads without being traced). Unequivocal symptoms (e.g. an extreme case is Ebola) and willingness to report them are requirements for a TETRIS strategy to work.

We developed an algorithm that for a given N-day long 3rd phase finds isolation levels for healthy <60 and vulnerable that minimizes final SARS-CoV-2 positive deaths. What happens when the N-day long 3rd phase ends? Some have proposed keeping isolation restrictions until the vaccine arrives (or even forever). Another alternative is to follow with a 4th phase with different isolation levels (same levels would be just a longer 3rd phase). In this case, final deaths will be lower or equal than with a uniform isolations phase but multiple solutions exist. To disambiguate, an additional minimizing criterion must be introduced (e.g. minimizing cumulative daily isolation values). Of all criteria, we chose a simple one: returning to normal life (i.e. all isolations in 0.00 when the 3rd phase ends).

Various final deaths minimizing strategies (without a vaccine in the future) are shown in Table 8 (see Graphic 3 for typical results but with a vaccine).

**Table 8.** Final deaths per million (ICU/100K), isolation to vulnerable (with 0.94 maximum), and isolation to healthy <60 for various strategies (beginning on lockdown day) and followed by normal life.

| Metropolitan area | Stockholm | Madrid | Catalonia | Brussels | Paris* | London* |
| --- | --- | --- | --- | --- | --- | --- |
| Back in March 2020<br>N-day death<br>minimizing + NL | 1115<br>(10 ICU)<br>97-day<br>(0.941,<br>+0.17) | 1269<br>(23 ICU)<br>102-day<br>(0.941,<br>+0.16) | 1257<br>(15 ICU)<br>100-day<br>(0.941,<br>+0.07) | 1011<br>(10 ICU)<br>101-day<br>(0.941,<br>+0.23) | 1019<br>(9 ICU)<br>113-day<br>(0.941,<br>+0.18) | 923<br>(9 ICU)<br>104-day<br>(0.941,<br>+0.26) |
| Fitted + 90-day death<br>minimizing with 0.00<br>to healthy <60 +NL | 1579<br>(2 ICU)<br>(0.25,<br>0.00) | 2426<br>(14 ICU)<br>(0.37,<br>0.00) | 2325<br>(25 ICU)<br>(0.71,<br>0.00) | 1765<br>(8 ICU)<br>(0.35,<br>0.00) | 1911<br>(19 ICU)<br>(0.78,<br>0.00) | 1798<br>(16 ICU)<br>(0.60,<br>0.00) |
| Fitted + 90-day death<br>minimizing + NL | 1454<br>(2 ICU)<br>(0.941,<br>-1.98) | 1834<br>(15 ICU)<br>(0.941,<br>-0.84) | 1630<br>(34 ICU)<br>(0.941,<br>-0.43) | 1297<br>(9 ICU)<br>(0.941,<br>-0.96) | 1384<br>(28 ICU)<br>(0.941,<br>-0.35) | 1314<br>(17 ICU)<br>(0.941,<br>-0.50) |

“Back in March 2020 N-day death minimizing + NL” shows that back at the beginning of the epidemic high isolation on vulnerable (i.e. 0.94) and soft touch on healthy <60 (i.e. from +0.07 to +0.27) was the death minimizing strategy for most locations (i.e. avoiding infections overshooting by soft ceiling above HIT). These “would have been” predictions show how close to death minimizing was Dr. Tegnell’s proposal (i.e. schools open but stadiums closed).

“Fitted + 90-day death minimizing with 0.00 to healthy <60 + NL” shows results if the fitted strategy in each location was followed with a 90-day phase with isolation to healthy <60 forced to 0.00. All minimums were found with isolation to vulnerable lower than the maximum available (i.e. maximum available was 0.94). In this case, herd immunity cannot be reached by overshooting healthy <60 infections and so the algorithm finds deaths minimum <u>without</u> using the maximum isolation to vulnerable.

“Fitted + 90-day death minimizing + NL” shows results if fitted strategies were followed by a 90-day death minimizing phase without limiting healthy <60 isolations. Naturally, deaths minimums were lower or equal than if limiting isolation to healthy <60 to 0.00. And, as expected, all deaths minimums were higher than if a death minimizing strategy would have been applied from the beginning.

And shockingly for all locations, sub-optimal past strategies (i.e. the fitted) led them to a point where their death minimizing 90-day strategies have negative isolation to healthy <60 (e.g. coronavirus parties). In these cases, the algorithm finds the minimum by overshooting healthy <60 infections and using the maximum available isolation to vulnerable. Negative isolation values imply effective R higher than Ro since R = Ro * (1-Iso_abl[d]) (see Appendix 1).

This proves that mandatory restrictions on healthy <60 can be sub-optimal and lead to more final deaths of vulnerable individuals when returning to normal life.

What happens if a vaccine is available in the future? Minimization results for Madrid with a vaccine with 77% uniform efficacy, that can be inoculated in an ideal 1-day long campaign, are shown in Graphic 3: the reported daily deaths, the predicted wave in Mar 2021 for the tentative 180-day (0.65, 0.40) strategy, the vaccination on March 31, 2021 (i.e. the steep change in recovered population), the alternative strategies (green color) and the immunity level for the shorter alternative (others not shown).

As seen in Table 8, for the 90-day alternatives death minimizing (1834 per million) also occurs with isolation to healthy <60 in -0.84 (and isolation to vulnerable in 0.941). Since the vaccine is too distant in the future it does not affect a 90-day long strategy. And a longer 180-day strategy but with lower isolation available to vulnerable (i.e. 0.65) also favors virus spreading among healthy <60 with minimum reached using 0.03 isolation to healthy <60.

If long strategies with high isolations are available minimizing strategies will include waiting for the vaccine. Here, in a 180-day 3rd phase (coupled with 0.941 maximum isolation to vulnerable) the minimum (1545 per million) is found using the maximum available value for isolation to healthy <60 (i.e. +0.400), that is the green wave peaking on Apr 2021. Vaccine arrives just in time to avoid the second wave.

**Graphic 3.**
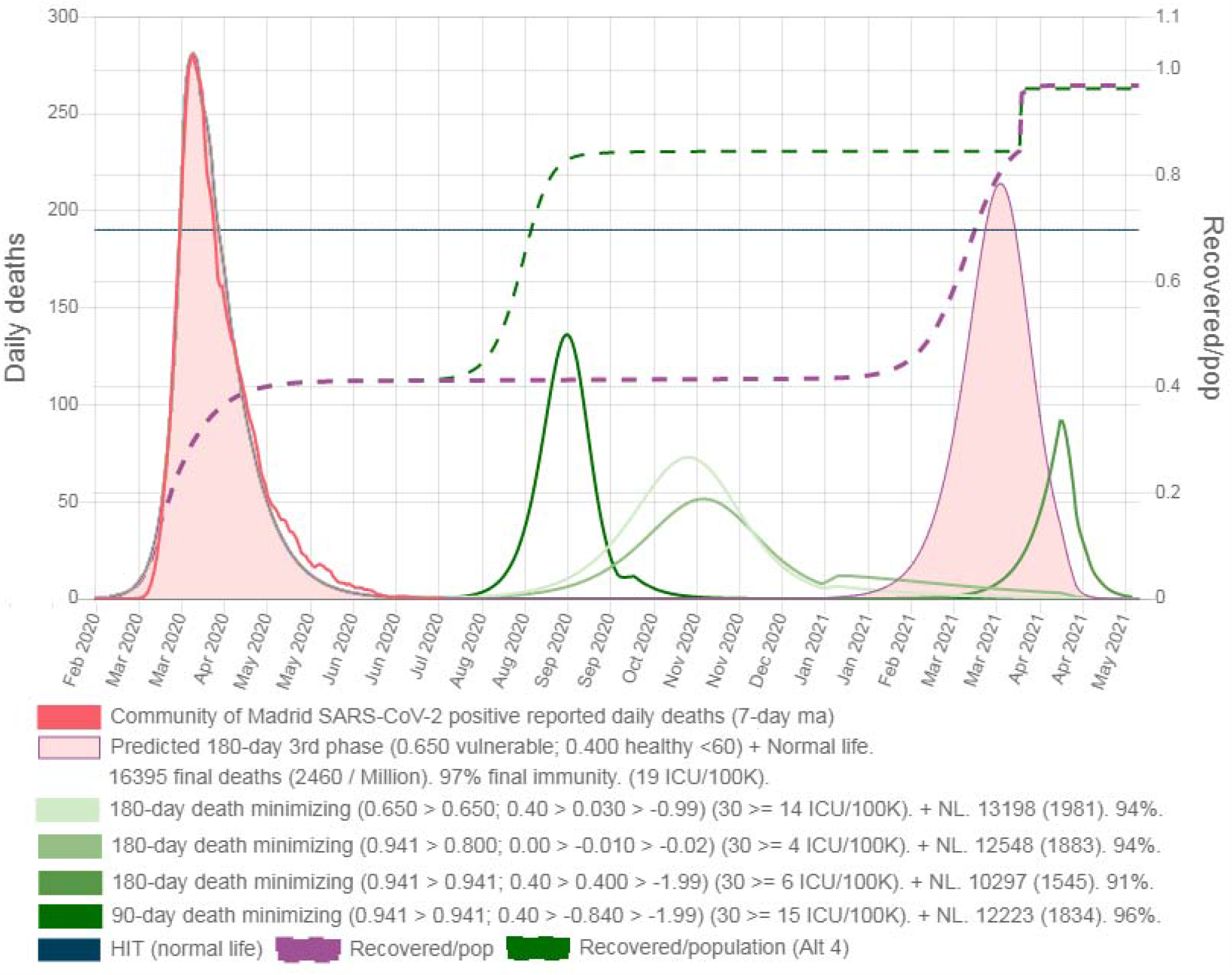
Death minimizing 3rd phase alternatives for Madrid (vaccination with 77% uniform efficacy on March 31, 2020).

Is it possible for vulnerable to isolate at 0.941 for 180-days? What if the vaccine does not arrive? After 6 months of harsh 0.94 isolation to vulnerable and 0.40 to healthy <60 another wave is waiting. It seems a costly bet. And as it was shown in Table 8 sub-optimal isolations strategies eliminate future death minimizing alternatives and thus result in more final deaths if the vaccine does not arrive.

### Final deaths for all vaccination dates

Assuming our 77% uniform efficacy vaccine (i.e. vEff) with a 1-day long vaccination campaign, we calculated final deaths for all possible vaccination dates for four strategies: Following the fitted strategy two 90-day death minimizing with 1) 0.94 and 2) 0.65 available isolation to vulnerable and 3) tentative 180-day (0.65, 0.40) aimed to wait for the vaccine, and 4) the “Back in March 2020 N-day death minimizing”. Results for Catalonia are shown in Graphic 4, and for all locations in Table 9.

**Table 9.** Final deaths per million for various death minimizing strategies for all possible dates of a vaccine with 77% efficacy.

| Metropolitan area | Stockholm | Madrid | Catalonia | Brussels | Paris* | London* |
| --- | --- | --- | --- | --- | --- | --- |
| Back in March 2020 no vaccine (NV) N-day death minimizing isolations asymptotic deaths/1M, ICUs, and (vaccine date for 99% asymptotic value) | 1110<br>97-day<br>(0.94,<br>0.17)<br>(12 ICU)<br>(May<br>20<br>2020) | 1269<br>102-day<br>(0.94,<br>0.16)<br>(15 ICU)<br>(May<br>5<br>2020) | 1248<br>100-day<br>(0.94,<br>0.07)<br>(15 ICU)<br>(May<br>6<br>2020) | 1011<br>101-day<br>(0.94,<br>0.23)<br>(10 ICU)<br>(May<br>10<br>2020) | 1019<br>113-day<br>(0.94,<br>0.18)<br>(11 ICU)<br>(May<br>11<br>2020) | 923<br>104-day<br>(0.94,<br>0.26)<br>(10 ICU)<br>(May<br>5<br>2020) |
| Asymptotic additional death/1M of Fitted + 180-day (0.65, 0.40) + NL vs. Fitted + 90-day (0.65) death minimizing + NL (first vaccine date that is positive) | +7<br>(0.65,<br>-0.62)<br>(Feb<br>11<br>2022) | +203<br>(0.65,<br>-0.46)<br>(Mar<br>13<br>2021) | +220<br>(0.65,<br>-0.10)<br>(Jan<br>29<br>2021) | +155<br>(0.65,<br>-0.51)<br>(May<br>17<br>2021) | +110<br>(0.65,<br>-0.07)<br>(Feb<br>20<br>2021) | +189<br>(0.65,<br>-0.22)<br>(Feb<br>5<br>2021) |
| Asymptotic additional death/1M of Fitted + 180-day (0.65, 0.40) + NL vs. Fitted + 90-day (0.95) death minimizing + NL (first vaccine date that is positive) | +15<br>(0.94,<br>-0.98)<br>(Jan<br>23<br>2022) | +650<br>(0.94,<br>-0.84)<br>(Feb<br>23<br>2021) | +865<br>(0.94,<br>-0.43)<br>(Dec<br>28<br>2020) | +514<br>(0.94,<br>-0.96)<br>(Apr<br>25<br>2021) | +663<br>(0.94,<br>-0.35)<br>(Jan<br>14<br>2021) | +586<br>(0.94,<br>-0.50)<br>(Jan<br>22<br>2021) |

**Graphic 4.**
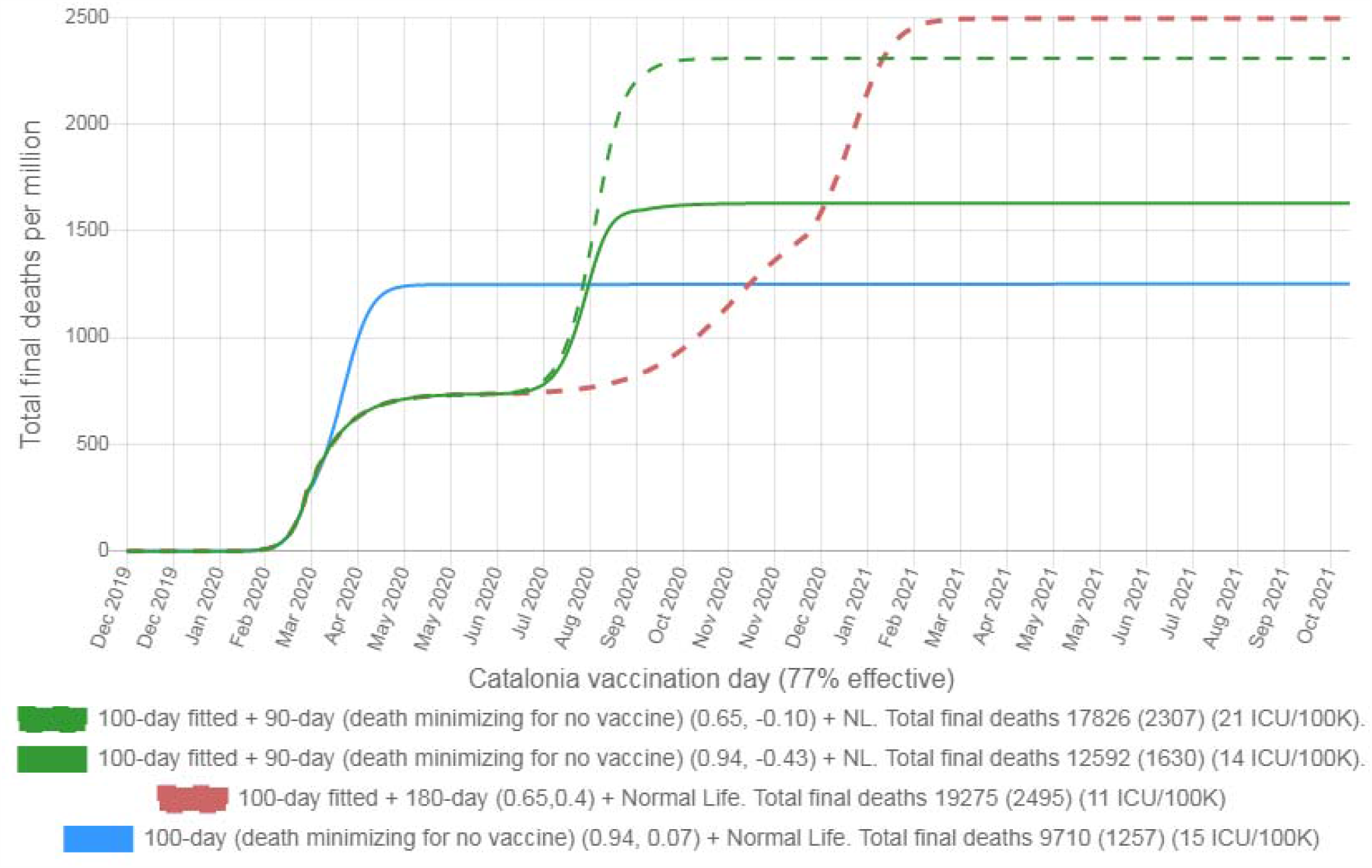
Final deaths per million in Catalonia for various isolation strategies for all vaccination dates with a 77 % effective and 1-day long campaign vaccine.

Since in Madrid any strategy that included the fitted strategy resulted in higher or equal deaths for every vaccination date than the “Back in March 2020 102-day death minimizing”, we switched to Catalonia (see Graphic 4) to show how the vaccine with 77% efficacy favors the 180-day (0.65, 0.4) strategy resulting in lower final deaths if vaccination occurs before November 2020 (i.e. when the dashed red line crosses the light blue solid one).

In Stockholm, final asymptotic deaths are similar for any strategy.

For the other locations, the ‘vaccine waiting’ strategy results in more deaths than the 90-day strategies if vaccination date is later than (Madrid: Feb 23 2021; Catalonia: Dec 28 2020; Brussels: Apr 25 2021; Paris*: Jan 14 2021; London*: Jan 22 2021;) and maximum isolation to vulnerable available for the 90-day strategies is 0.94 (and mostly less than 30 days later if it is 0.65).

**Graphic 5.1.**
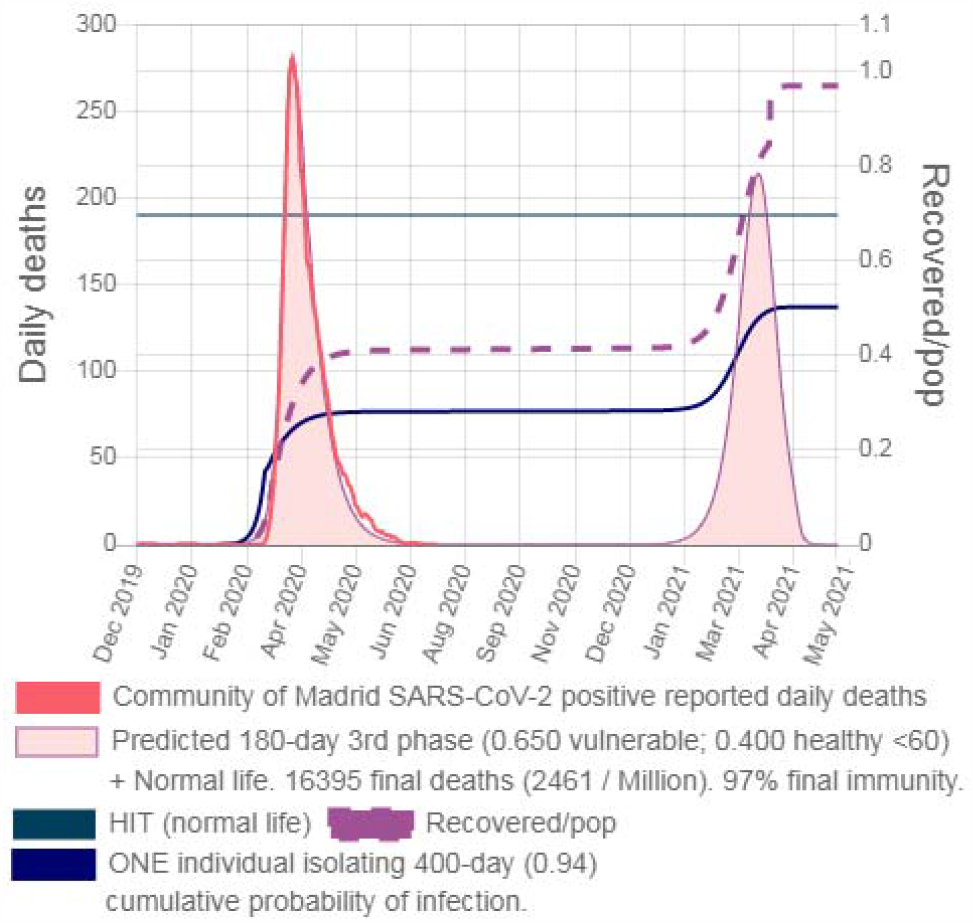
Madrid Fitted + 180-day (0.65, 0.40) + NL + vaccine with 77% efficacy on March 31 2021 1 individual forced to isolate for 400-day

**Graphic 5.2.**
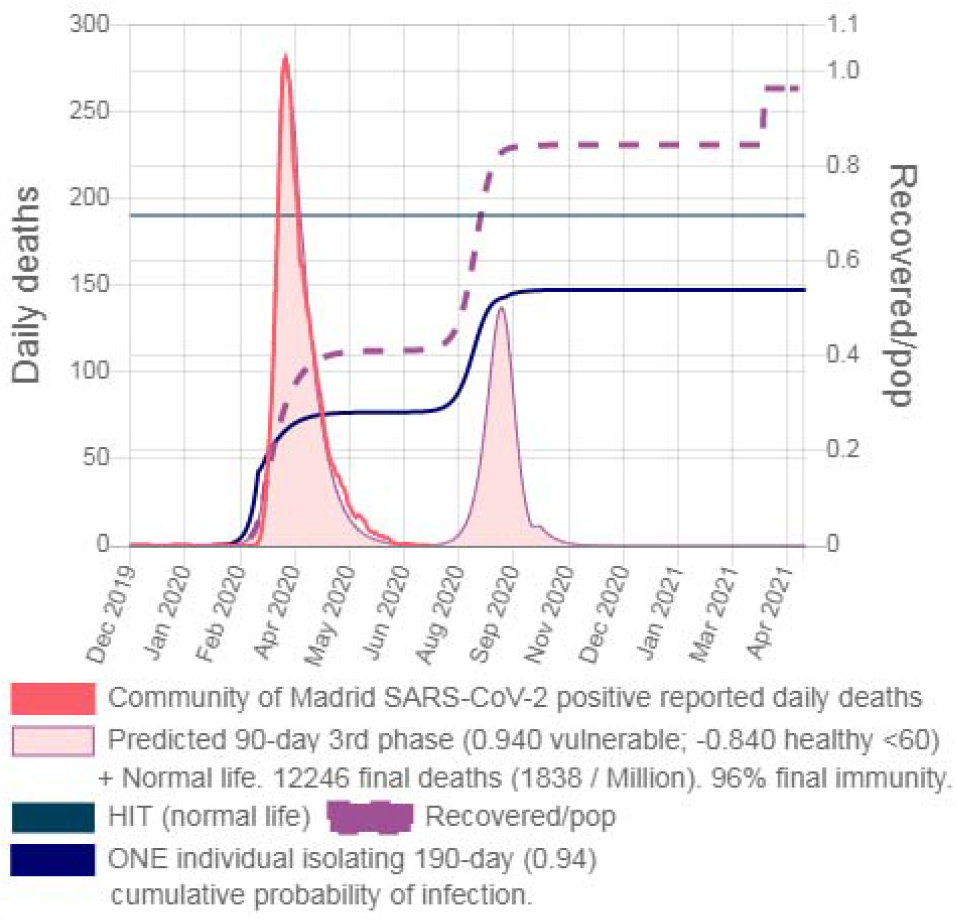
Madrid Fitted + 90-day death minimizing (0.94, -0.84) + NL + vaccine with 77% efficacy on March 31 2021 1 individual forced to isolate for 190-day

As for the implications for a single individual, he cannot avoid an increase in his probability of getting infected even if he continues to isolate at 0.94 until the vaccination date (see Graphic 5.1). On the other hand, he can cease to isolate after 190 days if the fitted strategy is followed by a 90-day (0.94, -0.84) strategy (Graphic 5.2). This example also shows that stratums isolations variations change the probability of one individual getting infected even though it continues to isolate at very high levels (e.g. 0.94)

**Graphic 6.**
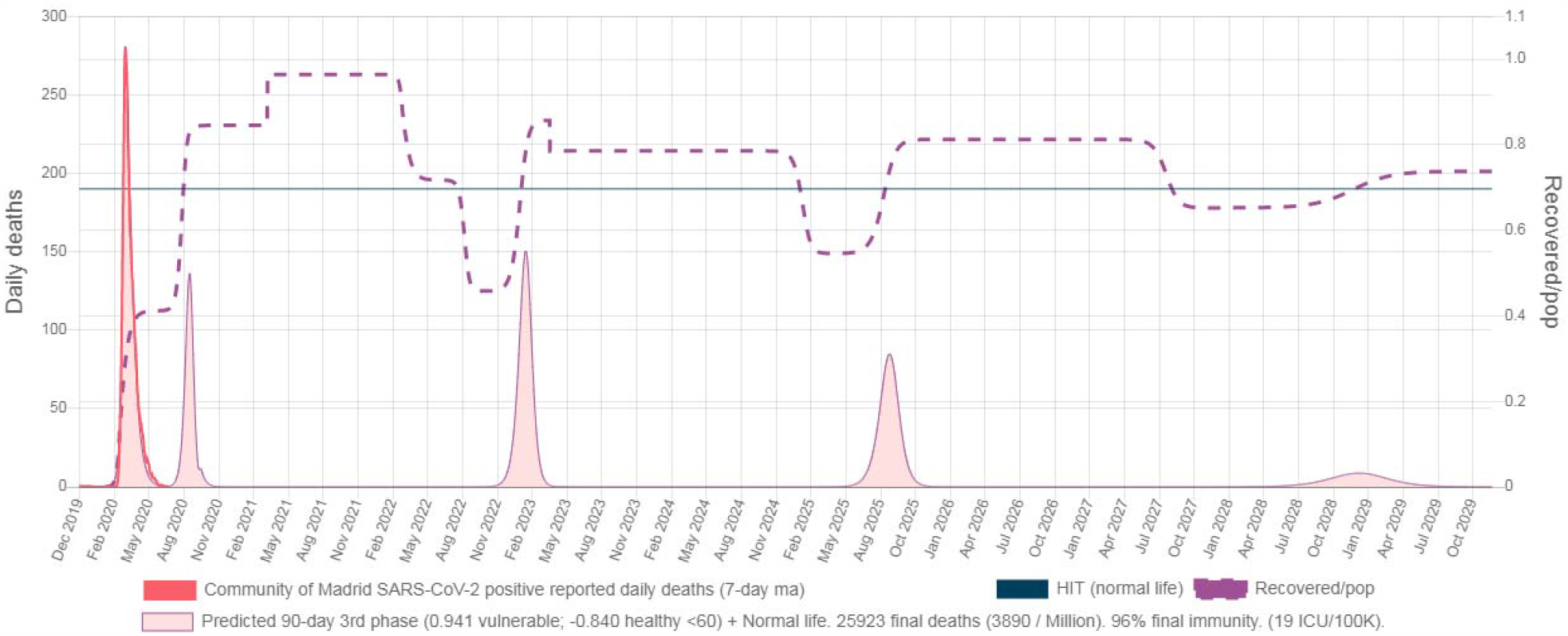
Effects of re-infections for a one-time vaccine followed by normal life and 60% of recovered and vaccinated losing immunity after 2 years.

In Graphic 6 we show a 10 years simulation if the vaccine can be used only once (e.g. immunity to the adenovirus vector lasting longer than to the SARS-CoV-2 virus) and assuming that 60% of recovered lose immunity (both from the infections and the vaccine) and become susceptible again after 2 years. The fitted strategy is followed by the 90-day death minimizing (0.941, -0.84) and then normal life, and the one-time vaccine with 77% efficacy is inoculated on March 31 2021.

### Isolation exemption insurance policy

State mandatory isolations have caused economic damages and since these enforced isolations were sub-optimal they involuntarily increased the risk of covid-19 disease-related damages (see additional deaths in Table 9) and led most locations to the point where future death minimizing strategies require negative isolation to healthy <60.

Back in March 2020 for every location (see Table 9) there was a 100-day long total death minimizing alternative (with low ICU occupation) that becomes asymptotic with lower values than 1269 deaths per million (and much lower person potential years live lost) if the vaccination date was later than May 2020 (that is May 2020 and it didn’t happen), and simultaneously mandatory isolations did continue to and still produce economic losses on daily basis. Therefore, we conclude that economic damages overcame covid-19 disease damages in all locations where governments kept enforcing mandatory isolation after June 2020.

What went wrong? The SARS-CoV-2 epidemic required complex risk assessment and governments are not the best equipped to do it (insurance companies are). We propose for this virus (and future viruses) an <u>isolation exemption insurance policy</u>. This should be evaluated both as a revealing thought experiment and a concrete suggestion.

In locations where mandatory isolations are imposed, any individual willing to be exempt from <u>complete</u> isolation during the epidemic must take out insurance to cover covid-19 disease damages to himself and also to third parties for spreading the virus (including years to live loss due to deaths or sequels and hospitalizations expenses). Why should a spreader pay an issue premium to cover covid-19 disease-related damages of third parties he will never see? Due to spreading a virus capable of causing disease causes damages.

Although an individual going to work during an epidemic may cause damages to third parties (and to himself), if the cost of covering the damages on third parties (or his own hospitalization expenses) is lower than his personal perceived cost of isolating he might decide to go out anyway.

Going out even though you can cause damage and covering the risk may seem heartless, but that is exactly what happens every time you drive your car or take a plane. Complete isolation of an elderly individual not willing to suffer damages from your behavior should include living inside a concrete bunker to avoid your car ending up inside his living room or the plane falling and destroying his roof. In an epidemic, the damaging spreader cannot be identified, and damage claims due to spreaders not isolating must be made to a common fund, like freeway insurance that pays damages for an accident caused by the rubber band from an anonymous broken tire.

The cumulative probability of an individual getting infected on day d of the epidemic is:

~~~
P_ind [d] = P_ind [d-1] + (1-P_ind[d-1])/pop * Ro/Do *
                        * (I_abl[d-1] * (1 - ind_abl_ci) + I_una[d-1] * (1 - ind_una_ci));
~~~

where I_abl[d-1] and I_una[d-1] are the amount of infected healthy <60 and vulnerable the prior day and ind_abl_ci and ind_abl_ci are the cross isolations of the individual with the healthy <60 and vulnerable groups and can be calculated as:

~~~
ind_abl_ci = 1 - (1-Iso_abl[d])**(1/2) * (1-Iso_ind[d])**(1/2); ind_una_ci = 1 - (1-Iso_una[d])**(1/2) * (1-Iso_ind[d])**(1/2);
~~~

where Iso_abl[d] and Iso_una[d] are the average isolations of healthy <60 and vulnerable groups, and Iso_ind is the personal isolation of the individual. Death probability on that individual depends on his personal IFR:

~~~
P_ind [d] * IFR_ind;
~~~

Hence, an individual not wanting to isolate at all will increase his personal disease damage risk due to his decision and thus will see increased the issue premium covering his own risk. And, since virus spreading will increase, issue premiums covering third-party damage claims will also increase. Or not.

That is the point; It should be clear by now that the impact of healthy <60 isolating 0.00 instead of 0.40 not always increases covid-19 damages. Back in March 2020 (uncertainty apart) issue premiums to healthy <60 willing to isolate 0.00 would have been higher because death minimizing isolation to healthy <60 was positive for most locations. But now, issue premiums with current vaccine efficacy and vaccination date probabilities should be zero for most locations because healthy <60 not isolating minimizes deaths.

Beware that the exemption is from complete isolation (i.e. 1.00) so by definition no individual without insurance could claim covid-19 damages. Most likely governments would allow lower values of uninsured isolations and cover the difference.

ICUs scarcity at the beginning of the epidemic may enormously increase disease damages due to improper medical attention. But if allowed, an increase in ICU hospitalization prices fuels rapid ICU expansion, and that risk is eliminated. Most likely insurance will have exclusions, for example going out if you are symptomatic or entering nursing homes or non-covid-19 hospitals without proper safety measures.

The proposed insurance would also help individuals to assess the personal risk they are subject to (e.g. for different isolation brackets) by knowing the issue premium for personal damages. At some point, a waiver to the personal damages claim will be the option for vulnerable individuals wanting to isolate below some threshold.

For a given moment, issue premiums and perceived personal isolation cost of all individuals will lead to equilibrium and average isolations for vulnerable and healthy <60 will continue to drive the epidemic behavior. If more than 2 isolation exemption insurance brackets are available, then extending to N-stratum isolations may be useful to analyze the 2-stratum average isolations sensitivity to issue premiums variations.

The absence or existence of the isolation exemption insurance to cover third party damages for a particular contagious disease could be a justification of why an Ebola patient can be forced to stay in a hospital room or why the “bubble boy” cannot force population lockdowns to avoid catching the common cold.

What happens if a virus is so contagious (high Ro) and so damaging (high IFR_vul) to large proportions of the population (high P_vul) that no solution exists to minimize damages and no insurer pool can cover the risk of exempting isolations? That virus pretty much will be the end of normal life. SARS-CoV-2 is not.

## Conclusions

The SARS-CoV-2 virus infection mortality rate has such a steep difference between vulnerable and healthy <60 individuals that stratified isolations result in fewer final deaths. Our proposed 2-stratum SEIRS model is able to fit past daily deaths curves using common virus parameters across all locations and months long constant isolation values for vulnerable and healthy <60 disambiguated with the reported age serology ratio. Hence, we maintain that is suitable for predicting deaths curves (and to minimize them) even under the possibility of a vaccination campaign. The probability of covid-19 disease-related damages suffered and caused by a single individual choosing not to completely isolate can also be estimated, which may allow calculating the cost of covering those risks with an isolation exemption insurance policy and thus help to minimize final deaths and economic losses caused by this epidemic.

## Data Availability

All data is available in the main text or the supplementary materials.

https://www.sars2seir.com/paper-12-2020/

## Appendix I SEIRS model with 2-stratum isolation

The classic Susceptible -> Exposed -> Infected -> Recovered model is extremely simple and ordered formulas for day d are:

~~~
nE[d] = S[d-1]/pop * Ro/Do * I[d-1]; nI[d] = nE[d-Eo];
nR[d] = nI[d-Do]; S[d] = S[d-1] - nI[d];
I[d] = I[d-1] + nI[d] - nR[d];
R[d] = R[d-1] + nR[d];
~~~

where nE: new exposed (communitarian), I: infected, S: susceptible individuals, pop: population, Ro: reproduction number, Do: days of infectiousness, nI: new infected, nR: new recovered, and R: recovered. Each infected individual infects other Ro and new infected grow until S[d]/pop * Ro < 1 or equivalent until (1-S[d]/pop) > 1-1/Ro, this value is called Herd Immunity Threshold (HIT). If infectiousness decays exponentially, new recovered can be expressed as:

~~~
nR [d] = I[d-1] / Do;
~~~

resulting in the same Ro (i.e. still each infected individual infects other Ro) but with a longer tail of infections (e.g. for Do=10 on the first 10 days of infectiousness only 61% of contagions occur). New deaths formula is:

~~~
nD [d] = IFR * nR [d];
~~~

where IFR is the infection fatality rate. If quarantine isolations are in place the probability of getting exposed is reduced and new exposed are:

~~~
nE[d] = S[d-1]/pop * Ro/Do * I[d-1] * (1 - Iso [d-1]);
~~~

where Iso is the isolation level. Ro*(1 - Iso) is known as effective R. To start the iteration all values in 0 except for:

~~~
S [0] = So * pop;
R [0] = (1- So) *pop;
~~~

where So is the proportion of susceptible population. To introduce non-communitarian exposed on S_day, between communitarian new exposition and susceptible calculations:

~~~
nE [S_day] = nE [S_day] + I_dayS;
~~~

where I_dayS are non communitarian new exposed on S_day (e.g. plane arrives). With S_day > 0 and I_dayS > S[S_day-1]. Intensive care unit (ICU) hospitalizations can be estimated with:

~~~
ICU_h[d] = ICU_pd * sum(nD[(d - ICU_dur) .. d])
~~~

where ICU_pd: average days of ICU hospitalizations per death, and ICU_dur: average ICU hospitalization period for all patients, and sum(nD) are the sum of new deaths during the next ICU_dur days. If after a period of time a proportion of recovered loss immunity, then:

~~~
nS [d] = nR[d-T1] * PRLI;
S[d] = S[d-1] - nI[d] + nS [d];
R[d] = R[d-1] + nR[d] - nS [d];
~~~

where nS[d]: new susceptible; T1: average period for losing immunity; and PRLI: proportion of recovered losing immunity (this is what is usually known as the SEIRS model). If it is relevant modeling PRLI as a function of period t,

~~~
nS [d] = sumAT(nR[d-t] * PRLI [t]);
~~~

where sumAT: sum for all values of t.

If immunity is not lost and a 2nd competing strain with complete cross immunity appears, then:

~~~
nE2[d] = S[d-1]/pop * Ro2/Do2 * I2[d-1] * (1 – Iso2 [d-1]);
nI2[d] = nE2[d-Eo2];
nR2[d] = nI2[d-Do2];
S[d] = S[d-1] - nI[d] – nI2[d];
I2[d] = I2[d-1] + nI2[d] – nR2[d];
R2[d] = R2[d-1] + nR2[d];
~~~

where the 2nd strain obtains and draws new exposed from the same susceptible compartment. If Ro2 > Ro and recovered are far from HIT, then I2 >> I and the 2nd strain eventually overcomes the 1st. Deaths from the 2nd strain:

~~~
nD2 [d] = IFR2 * nR2 [d];
~~~

where IFR2: infection fatality rate of the 2nd strain. In what scenario can Iso2 be different than Iso? For example, if the 2nd strain produces fewer symptoms and symptomatic stay home. Usually, fewer symptoms mean less mortality (i.e IFR2 < IFR), and thus symptomatic staying home results in less final deaths.

If recovered from a 2nd strain with no cross immunity (i.e. almost another virus) become susceptible to the 1st strain but with IFR3 <> IFR (e.g. IFR3 > IFR in Dengue) then a third computation must be performed for the spreading of the 1st strain:

~~~
nS3[d] = nR2[d-T3] * PRLI3;
nE3[d] = S3[d-1]/pop * Ro/Do * ((I[d-1]+ I3[d-1]) * (1 – Iso [d-1]);
nE[d] = S[d-1]/pop * Ro/Do * ((I[d-1]+ I3[d-1]) * (1 – Iso [d-1]);
nI3[d] = nE3[d-Eo];
nR3[d] = nI3[d-Do];
S[d] = S[d-1] - nI[d] - nS3[d];
S3[d] = S3[d-1] – nI3[d] + nS3[d];
I3[d] = I3[d-1] + nI3[d];
R3[d] = R3[d-1] + nR3[d];
~~~

and

~~~
nD3 [d] = IFR3 * nR3 [d];
~~~

Where nS3: new susceptible to the 1st strain with IFR3; nR2: recovered from the 2nd strain; T3: average period from recovered of 2nd strain to becoming susceptible with IFR3 to the 1st strain; PRLI3: proportion of recovered from 2nd strain that becomes susceptible with IFR3 to the 1st strain. This last example shows how highly complex scenarios can be easily modeled by adding simple algebraic operations to the main iteration.

### Simple extension for 2-stratum isolation

To extend for isolation stratification the population is separated in two different groups (e.g. A for able to isolate from vulnerable and U for unable to isolate from vulnerable) and the new exposed for day d are calculated with:

~~~
nE_abl[d] = S_abl[d-1]/pop*Ro/Do * (I_abl[d-1]*(1-Iso_abl[d]) + I_una[d-1]*(1-auci[d]));
nE_una[d] = S_una[d-1]/pop*Ro/Do * (I_una[d-1]*(1-Iso_una[d]) + I_abl[d-1]*(1-auci[d]));
~~~

where nE_abl: communitarian new exposed of the A group, S_abl: susceptible of A, I_abl: infected of A, Iso_abl: restriction level between A individuals, I_una: infected of U, nE_una: communitarian new exposed of U, S_una: susceptible of U, I_una: infected of U, Iso_una: restriction level between A individuals, and auci: cross isolation between A and U isolation groups. A simple hypothesis for auci used here is:

~~~
auci[d] = 1-Math.sqrt(1-Iso_abl[d]) * Math.sqrt(1-Iso_una[d]);
~~~

New exposed of the A group (nE_abl) are susceptible of the A group that get exposed from the currently infected of the A group at Ro/Do * (1-Iso_abl) daily rate and from the currently infected of the U group at Ro/Do * (1-auci) daily rate. The analogous holds for new exposed for the U group (nE_una).

Susceptible, infected, and recovered of the two groups do not interact in another way and classic SEIRS applies. If uniform non-susceptibility is used, to start the simulation:

~~~
S_abl [0] = So * pop * P_abl;
S_una [0] = So * pop * (1-P_abl);
R_abl [0] = (1-So) * pop * P_abl;
R_una [0] = (1-So) * pop * (1 - P_abl);
where P_abl: proportion of the population that belongs to group A, calculated here as:
P_abl = 1-(P_vul + P_non_vul_una);
~~~

where P_vul: proportion of vulnerable population (i.e. a higher IFR applies to them), P_non_vul_una: proportion of population non-vulnerable unable to isolate from vulnerable (e.g. 7%, living in the same household). P_vul is calculated as:

~~~
P_vul = P_o60 + (1-P_o60) * P_vul_u60;
~~~

where P_o60: proportion of the population older than 60 years old (e.g. 29% in Europe) and P_vul_u60: proportion of younger than 60 years population that is vulnerable (e.g. 3.42%, see Appendix II for parameter estimations discussion). For Europe:

~~~
P_vul = 0.29 + (1 – 0.29) * 0.0342 = 0.3143
P_abl = 1 - (0.3143 + 0.07) = 0.3843
~~~

To introduce uniform non communitarian exposures en S_day, between communitarian new exposition and susceptible calculations:

~~~
nE_abl [S_day] = nE_abl [S_day] + I_dayS * P_abl;
nE_una [S_day] = nE_abl [S_day] + I_dayS * (1-P_abl);
~~~

And finally to extend to mortality stratification, since some non vulnerable individuals are isolated with vulnerable new deaths from each isolation group are calculated as follows:

~~~
nD_vul [d] = IFR_vul * nR_U[d-(RtoD-Do)] * P_vul/(P_vul + P_non_vul_una);
nD_non_vul [d] = IFR_non_vul *
       * (nR_A[d-(RtoD-Do)] + nR_U[d-(RtoD-Do)] / (P_vul / P_non_vul_una +1));
~~~

where nD_vul: new deaths of vulnerable individuals, IFR_vul: infection fatality rate for vulnerable individuals (e.g. 1.06%), RtoD: average period between “recovered” and death (e.g. 10 days), and IFR_non_vul: IFR for non vulnerable individuals (e.g. 0.01%). And to calculate new recovered <60 and >60:

~~~
nR_o60[d] = nR_una[d] * P_o60 / (P_o60 + (1-P_o60) * P_vul_u60 + P_non_vul_una);
nR_u60[d] = nR_abl[d] + nR_una[d] * ((1-P_o60) * P_vul_u60 + P_non_vul_una) / (P_o60 + (1-P_o60) *
        * P_vul_u60 + P_non_vul_una);
~~~

For a uniform vEff effective vaccination event occurring on vDay, after calculating for day d:

~~~
nR_vacc_abl [d] = S_abl[d] * vEff;
nR_vacc_una[d] = S_una[d] * vEff;
R_abl[d] = R_abl[d] + nR_vacc_abl [d];
R_una[d] = R_una[d] + nR_vacc_una[d];
S_abl[d] = S_abl[d] - nR_vacc_abl[d];
S_una[d] = S_una[d] - nR_vacc_una[d];
~~~

Exact modeling of a non uniform vaccination event (e.g. not all vulnerable can be vaccinated) would require healthy <60 unable to isolate with vulnerable (P_non_vul_una) to be calculated separately (it will still be a 2-stratum isolation though). New susceptible losing immunity from infection and from vaccination are:

~~~
nS_una [d] = nR_una [d-T1] * PRLI + nR_vacc_una [d-T_vacc] * PRLI_vacc;
nS_abl [d] = nR_abl [d-T1] * PRLI + nR_vacc_abl [d-T_vacc] * PRLI_vacc;
~~~

where T_vacc and PRLI_vacc are the average period and proportion of recovered losing vaccine immunity.

If one single individual choose to isolate different than any average the probability of getting infected can be calculated with:

~~~
P_ind [d] = P_ind [d-1] + (1-P_ind[d-1])/pop * Ro/Do *
          * (I_abl[d-1] * (1 - ind_abl_ci) + I_una[d-1] * (1 - ind_una_ci));
~~~

where P_ind: cumulative probability of getting infected on day d; ind_abl_ci: individual and able cross isolation, ind_abl_ci: individual and unable cross isolation, and both are calculated:

~~~
ind_abl_ci = 1-Math.sqrt(1-Iso_abl[d]) * Math.sqrt(1-Iso_ind[d]);
ind_una_ci = 1-Math.sqrt(1-Iso_una[d]) * Math.sqrt(1-Iso_ind[d]);
~~~

where Iso_ind: individual isolation.

Note that shall the single individual isolate at the same level that vulnerable (i.e. Iso_una) then ind_una_ci = Iso_una, and ind_abl_ci = auci (i.e. vulnerable and healthy <60 cross isolation) and the following holds:

~~~
nE_una [d] = (1-P_ind[d-1])* S_una[0]/pop * Ro/Do *
* (I_abl[d-1] * (1 - auci) + I_una[d-1] * (1 – Iso_una[d-1]));
~~~

The extension of the classic SEIRS model is then a small set of simple iterative instructions. The exact same model used through this paper (1) has been made available online to the sole effect of understanding and verifying its content.

## Appendix II Parameters discussion and immune level estimation sensitivity

Immunity value estimation sensitivity analysis to parameter variation was performed changing each variable to Min and Max values and fitting daily deaths curve again (total, peak 7-day moving average, reported serology ratio) allowing variations of infected on spread day, isolation to healthy <60, and isolation to vulnerable.

### Common virus

Sensitivity analysis to common virus parameters variations is shown in Table A.1.

**Table A.1.** Madrid immunity level estimation (Recovered/pop on July 2020) sensitivity for virus parameters.

| Variable name | Variable values | Immunity value | Sensitivity (multiplier) |
| --- | --- | --- | --- |
|  | Point (Min : Max) | Point (Min : Max) |  |
| Ro | 3.3 (2.5 : 4.0) | 41% (41% : 41%) | ≈ 0 |
| Do | 2 (1.33 : 5) | 41% (41% : 41%) | ≈ 0 |
| Eo | 5 (4 : 6) | 41% (41% : 41%) | ≈ 0 |
| RtoD | 11 (9 : 13) | 41% (41% : 41%) | ≈ 0 |
| So | 1.00 (0.9 : NA) | 41% (51% : NA) | -2.44 |
| IFR_vul | 0.0092 (0.0077 : 0.0104) | 41% (48% : 37%) | -0.85 |
| IFR_non_vul | 0.000035 (0.000029 : 0.000041) | 41% (41% : 41%) | ≈ 0 |
| P_vul_u60 | 0.0342 (0.0274 : 0.0401) | 41% (42% : 41%) | -0.06 |
| P_non_vul_una | 0.07 (0.05 : 0.09) | 41% (41% : 41%) | ≈ 0 |
| T1 | 180 (150 : 210) | 41% (41% : 41%) | ≈ 0 |
| PRLI | 0.00 (NA : 0.50) | 41% (41% : 41%) | ≈ 0 |
| Ma | 7 (1 : 14) | 41% (41% : 41%) | ≈ 0 |
| A_o60 | 0.21 (0.17 : 0.25) | 41% (42% : 41%) | -0.06 |
| A_u60 | 0.52 (0.42 : 0.62) | 41% (40% : 43%) | +0.21 |
| TAK | 0.35 (0.29 : 0.42) | 41% (41% : 42%) | +0.07 |
| APTP | 17 (13 : 21) | 41% (43% : 40%) | -0.17 |
| ICU_pd | 0.45 | NA | NA |
| ICU_h_dur | 10 days | NA | NA |
| vEff | 0.77 | NA | NA |

#### Ro = 3.3 (sensitivity ≈ 0)

Reproduction number for normal life. Should be estimated before restriction/lockdowns are in place. Here we assume is constant among metropolis. It does not affect intermediate immunity level estimations but determines the immunity level necessary to return to normal life HIT=1-1/Ro. Large variations in Ro estimations (e.g. 2.5 to 4.0) do not produce large variations in HIT (i.e. 60% to 75%).

#### Do = 2 days (sensitivity ≈ 0)

Average infectiousness period. See 2 day Do (infectiousness) section for point estimation.

#### Eo = 5 days (sensitivity ≈ 0)

Exposure period. See 2 day Do (infectiousness) section for point estimation.

#### RtoD = 11 days (sensitivity ≈ 0)

Recovered to death period. Estimated by fitting daily deaths curves for all locations near lockdown day.

#### So = 1.00 (sensitivity -2.44)

Proportion of population that is susceptible, we assume 1 (novel virus). Also back in March 2020 when isolations have not had effect yet SARS-CoV-2 positive deaths had a steep increase in all analyzed locations pointing to high proportion of susceptible.

#### IFR_vul = 0.0092 (sensitivity -0.85)

Infection Fatality Rate for vulnerable individuals (probability of dying because of covid-19).

IFR_vul is an average value for >60 individuals (and healthy <60 with underlying diseases), stratifying an arbitrary amount of age segments to adjust a particular location for different age distributions of >60 population can also be done.

Beware that since the model predicts SARS-CoV-2 positive deaths including deaths of other causes testing positive by chance, to compare other IFR estimations for >60 with IFR_vul they should be decreased 13.2% (or equivalently IFR_vul increased to 1.06%). See section 15% of SARS-CoV-2 positive deaths are “with” the virus in Appendix III - Secondary findings.

And since most IFR estimations do not account for undetected T cell immune response to be compared with IFR_vul they should be decreased 8%. See section Asymptomatic: 52% of <60 and 21% of >60 in Appendix III - Secondary findings.

In (16) estimated IFR (95% CI) for older >60 not living in nursing homes was 1.71% (1.28%-2.58%). Since all >60 are vulnerable and decreasing the IFR estimation 8% (for undetected T cells) and 13.2% (for testing positive by chance) results in 1.38% (1.03% - 2.08%) (95% CI), then IFR_vul is still below the lower bound of the CI reported in (16). Why do we use this lower value of IFR_vul anyway? We are estimating immunity levels and predicting epidemic behavior for real and we think that IFR_vul is currently overestimated because of vanishing antibody titer levels (33).

The wide range of IFR estimations reported in (23) shows the difficulties of using absolute values of seroprevalence studies to estimate IFR. Adding competing SARS-CoV-2 strains with different IFR_vul and infectivity (see Early D614 like strain wave in Asia: A hypothesis in Appendix III) would result in different IFR average estimations depending on the prevalence of each strain. Besides age and underlying diseases, ethnicity (16) and blood type (39) among others have also been reported to affect IFR. Medical treatments of covid-19 are also constantly improving (e.g. Dexamethasone results reported in (40)) but they may not always be used homogeneously across locations.

Lower real values of IFR_vul than the used here (i.e. 0.92%) would mean that our immunity levels estimations are a lower bound of real values. But beware that death minimizing isolation strategies resulting in lower values of isolation to healthy <60 than isolation to vulnerable do not arise from any particular value of IFR_vul, but because of the unequivocal fact that IFR_non_vul is much lower than IFR_vul.

#### IFR_non_vul = 0.000035 (sensitivity ≈ 0)

IFR for healthy <60 (probability of dying because of covid-19).

In (28) were reported 89 no illness deaths of <60 and 414 with pending investigation. We have fitted NYC and the model predicts 772 SARS-CoV-2 positive deaths for healthy <60, so even counting all pending investigation as no illness <60 deaths predicted deaths are higher than reported ones. Hence IFR_non_vul used here can be considered an upper bound of the real IFR for healthy <60.

#### APTP = 17 (sensitivity -0.17)

Average positive testing period.

In (33) the median duration of viral shedding was 19 days for asymptomatic and 14 days for symptomatic. Given the 0.52 proportion for asymptomatic in <60 and 0.21 in >60 for all practical P_o60 proportions (i.e. 0.1 to 0.3) we assume APTP = 17. Different APTP values for vulnerable and healthy <60 could also be used.

#### P_vul_u60 = 0.0342 (sensitivity -0.06)

Proportion of <60 population that is vulnerable. Using IFR ranges estimated in (16) and Indiana population pyramid, IFR for <60 is estimated at 0.046%. Formula for IFR <60 is

~~~
0.00046 = P_vul_u60 * IFR_vul_sc2p + (1-P_vul_u60) * IFR_non_vul_sc2p
~~~

with P_vul_u60 = 0.0342, if IFR_vul_sc2p (i.e. 0.0106) and IFR_non_vul_sc2p (i.e. 0.0001) as defined in section: 15% of SARS-CoV-2 positive deaths are “with” the virus.

#### P_non_vul_una = 0.07 (sensitivity ≈ 0)

Proportion of population that is <60 and cannot isolate from vulnerable individuals. Average guessing.

#### T1 = 180 (sensitivity ≈ 0)

Period between recovered and losing immunity (days). No estimations were available.

#### PRLI = 0.00 (sensitivity ≈ 0)

Proportion of recovered losing immunity after T1. No estimations were available.

#### A_o60 = 0.21 (sensitivity -0.06)

Proportion of >60 that are asymptomatic. Selected to fit (8) and in line with (27). See Asymptomatic section in Appendix III.

#### A_u60 = 0.52 (sensitivity +0.21)

Proportion of <60 that are asymptomatic. Selected to fit (8) and in line with (27). See Asymptomatic section in Appendix III.

#### TAK = 0.35 (sensitivity +0.07)

Proportion of asymptomatic with T cell response but without IgG response (15). See Asymptomatic section in Appendix III.

#### Ma = 7 days (sensitivity ≈ 0)

Moving average filter.

#### ICU_pd = 0.45 (sensitivity not applicable)

ICU admissions per SARS-CoV-2 positive death. (30)

ICU_h_dur = 10 (sensitivity not applicable) Average ICU hospitalizations duration. (2)

#### vEff = 0.77 (sensitivity not applicable)

Vaccine efficacy. Target vaccine efficacies are 60 % (31)(35) and we assume for vEff the highest success criteria of 77% reported in (31).

#### Location-specific

Sensitivity analysis to location-specific parameters variations is shown in Table A.2.

**A.2.**
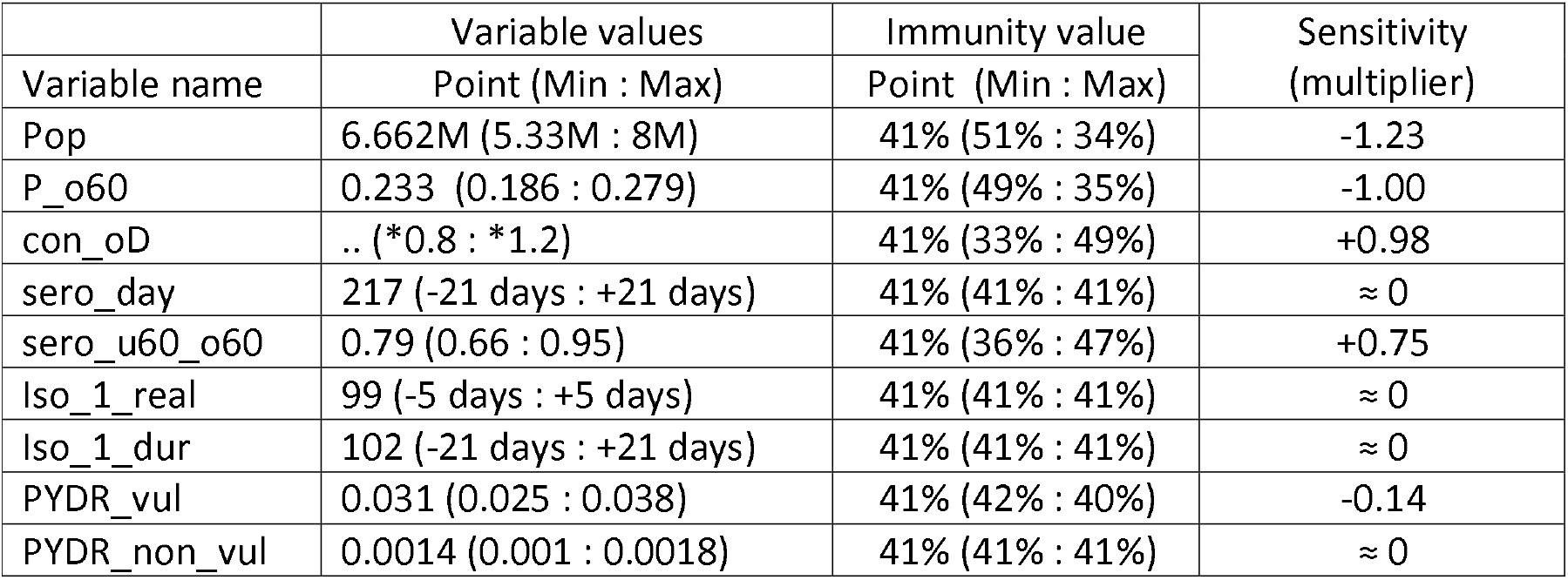
Madrid immunity level estimation (Recovered/pop) sensitivity for location parameters.

Each variable was changed to Min and Max values and daily deaths curve fitting was performed (total, peak 7-day moving average, reported serology ratio) allowing variations of infected on spread day, isolation to healthy <60, and isolation to vulnerable.

#### Pop (sensitivity -1.23)

Population of metropolitan area. The location must be a single conglomerate to avoid separated multiple waves (i.e. statewide or countrywide simulations will not produce meaningful predictions).

#### P_o60 (sensitivity -1.00)

Proportion of population of the metropolitan area older than 60 years. Metropolitan areas cannot be approximated with countries population pyramids because proportion of >60 population is usually lower in metropolis.

#### con_oD (sensitivity +0.98)

Reported daily deaths. SARS-CoV-2 positive deaths and recorded in the exact deceased date.

Recording the exact date will change past reported daily deaths (i.e. usually deaths are not reported the same day they occurred) but sensitive information may be lost if the exact day of deceased is not recorded.

It should also include testing deceased post-mortem if a near-death test (e.g. PCR) was not performed (most countries do it that way). Some locations (e.g. Singapore) might have instructed medical doctors to discriminate dying “with” or “because” of the virus and sensitive epidemiological information can be lost if dying “with” the virus is not recorded too. It is possible to match the “predicted reported deaths” produced by the model with the “with the virus” reported deaths and much harder to match it if a “heterogeneous per-doctor because of the virus” criterion is used for reporting. Though, if the chance of psychosis is considered, reporting “because” may make more sense.

A metropolitan area should be fitted as one single location. For example, Paris City departments are 75, 92, 93, 94 and Greater Paris also includes 95, 78, 91, and 77. Single cities inside conglomerates may have daily deaths of outsiders recorded in their hospitals and hence result in immune level overestimation. Also, deaths from all districts must be included to avoid immune level underestimation.

In some locations reported daily deaths might be available only from hospitals since total deaths for locations are usually well known daily deaths can be corrected by using the con_oD_scale variable in the

*.js location file (e.g. we did it with Greater London con_oD_scale = 1.11).

#### PYDR_vul = 0.031 (sensitivity -0.14)

Population yearly death rate for vulnerable individuals (>60 natural). Using population yearly death rate by age published in (32) with the >60 population pyramid PYDR_vul = 0.031.

#### PYDR_non_vul = 0.0014 (sensitivity ≈ 0)

Population yearly death rate for non-vulnerable individuals. Using population yearly death rate by age published in (32) with the population pyramid, yearly death rate for <60 is 0.00241. But since the <60 population includes a proportion of vulnerable (i.e. P_vul_u60 = 0.0342) we assume that:

~~~
0.00241 = P_vul_u60 * PYDR_vul + (1-P_vul_u60) * PYDR_non_vul
~~~

and hence we estimate PYDR_non_vul = 0.0014.

#### Iso_1_real (sensitivity ≈ 0)

Day of lockdown/restrictions beginning. Lockdowns beginnings are fuzzy, Iso_1_real is inferred from fitting daily deaths curves using uniform RtoD = 11 for all locations.

#### sero_u60_o60 (sensitivity +0.75)

<60 to >60 reported serology ratio.

The best seroprevalence studies are randomized and in theory should look for any kind of immune response that demonstrates exposure. Spain’s IgG antibody study (8) is the gold standard for randomization but an industrial T cell response test was not available at the time.

The reported serology ratio may change during time reflecting changes in individual behavior (e.g. healthy <60 summer behavior in London). But can also remain fairly constant like in Spain from May to June. The model used here supports multiple serology report dates.

If antibody titer levels in >60 individuals decrease slower than in <60 individuals then the reported serology ratio will be a lower bound of the real immune response and hence immunity level will be underestimated.

#### sero_day (sensitivity ≈ 0)

Day of reported serology levels.

#### Iso_1_dur (sensitivity ≈ 0)

Days of 1st phase isolation duration.

Large variations in reported serology ratio should be modeled with different phases to reflect the real isolations variations. Serology ratio at the end of the isolation phase cannot be used if was much lower at its beginning.

## Appendix III Secondary findings

### Asymptomatic: 52% of <60 and 21% of >60

By dividing <60 and >60 seroprevalence absolute values, serology tests inaccuracies are mostly canceled. But if the asymptomatic proportion of <60 is different than asymptomatic proportion of >60, then the age serology ratio cannot be estimated only with the <60 to >60 recovered ratio and the following should be used:

~~~
R_u60 / R_o60 * P_o60 / (1 - P_o60) / * (1 - A_u60 * TAK) / (1 - A_o60 * TAK)
~~~

where R_u60/R_o60 is the <60 and >60 recovered ratio, P_o60 is the proportion >60 in population, A_o60 and A_u60 are the proportion of >60 and <60 that are asymptomatic, and TAK is the proportion of asymptomatic with T cell response but without antibody detectable titer levels found in (15) (i.e. 0.35).

Missing IgG response in asymptomatic would not affect serology ratio if <60 and >60 have the same proportion of asymptomatic, but evidence points a different direction. In (27) reported median age in asymptomatic (37 years old) was lower than in symptomatic (56 years old).

We did not find age-stratified asymptomatic proportion reliable estimations (raw data in (8) may have it). But in order to fit the total asymptomatic to symptomatic ratio reported in the serology study in Spain (i.e. 0.51 = 34% / 66%), values A_o60 = 0.21 and A_u60 = 0.52 were necessary.

With those asymptomatic proportions (1 - A_u60 * TAK) / (1 - A_o60 * TAK) = 0.88. Implicit average proportion of asymptomatic in Madrid is 44.49% (in line with the consensus of 45% reported in 28).

### 15% of SARS-CoV-2 positive deaths are “with” the virus

If the criterion to report daily deaths is counting any death testing SARS-CoV-2 positive (as most locations do) the new deaths (nD) formula detailed in Appendix I cannot be used to predict them and the following should be used instead:

~~~
nD_vul[d] = IFR_vul_sc2p * nR_una[d-RtoD] * P_vul / (P_vul + P_non_vul_una);
~~~

where IFR_vul_sc2p: probability of dying testing sar-cov-2 positive (after being infected with SARS-CoV-2) and can be calculated as:

~~~
IFR_vul_sc2p = IFR_vul * (1-NDR_vul) + NDR_vul * (1-IFR_vul) + IFR_vul * NDR_vul;
~~~

where NDR_vul: probability of dying of other causes than sar-cov-2 testing sar-cov-2 positive (after being infected with SARS-CoV-2) and can be estimated as:

~~~
NDR_vul = (1+PYDR_vul) ** (APTP/365) - 1;
~~~

where PYDR_vul: population yearly death rate of other causes than SARS-CoV-2 infection for vulnerable individuals, ** indicates power operation, and APTP: average period of testing positive (in days) after being

infected to the virus (e.g. 17). IFR_vul_sc2p formula holds because NDR_vul and IFR_vul correspond to independent events. Being SARS-CoV-2 a novel virus and assuming PYDR_vul have not changed in 2020 (e.g. lockdowns have not markedly changed other than SARS-CoV-2 death rates), then prior to 2019 estimations can be used for PYDR_vul (e.g. 0.031 for >60 in 2017).

The analogous holds for nD_non_vul, but since <60 includes a proportion of vulnerable (i.e. P_vul_u60) then PYDR_non_vul (i.e. 0.0014) cannot be estimated by directly using <60 population yearly death rate (i.e. 0.00241). See Appendix II for details.

**Graphic A3.1.1.**
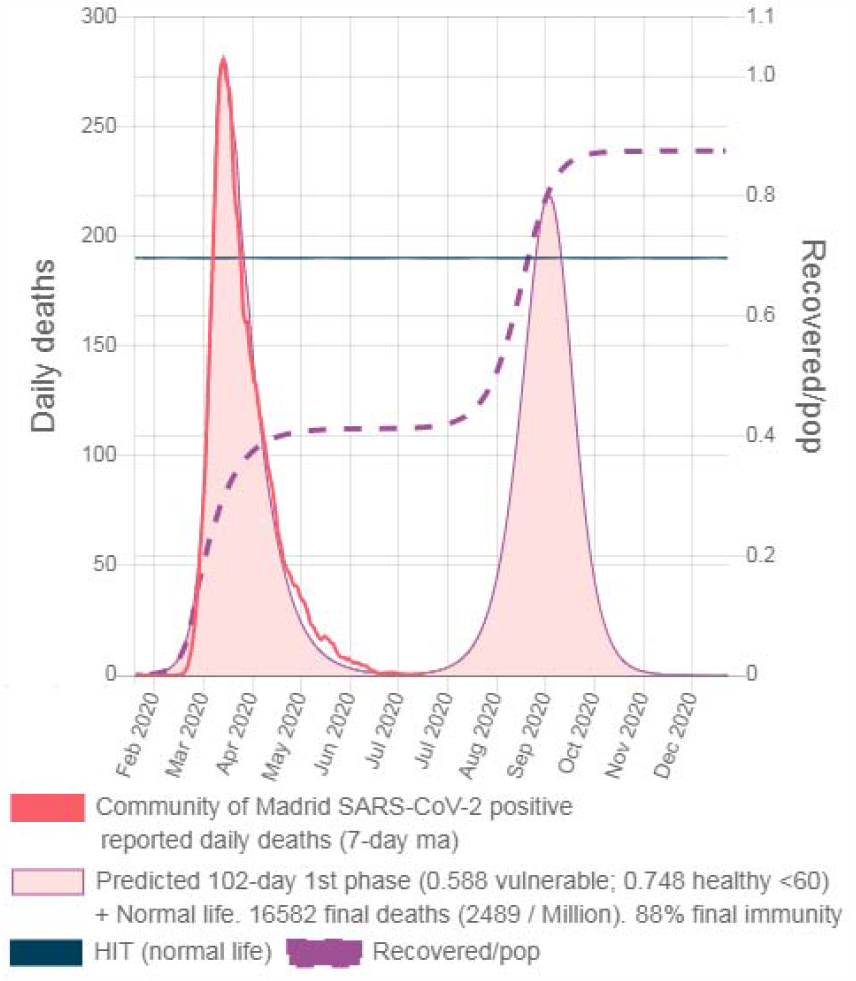
Madrid predicted SARS-CoV-2 positive deaths for SARS-CoV-2 virus (IFR_vul = 0.92%, IFR_non_vul = 0.0035%)

**Graphic A3.1.2.**
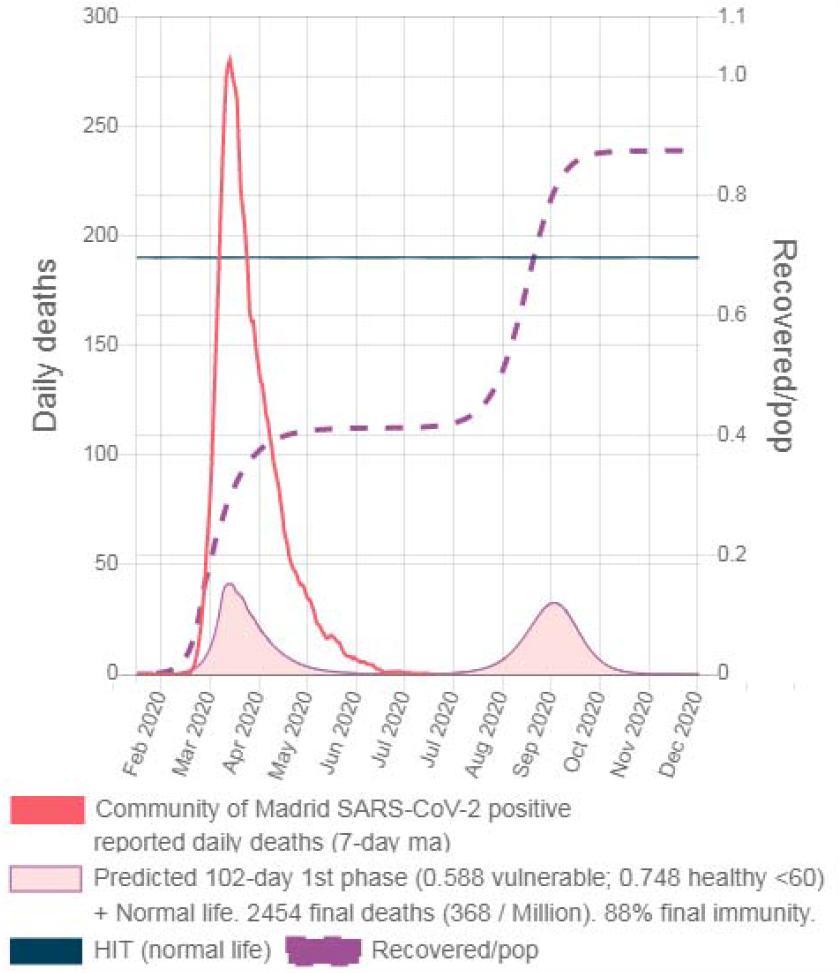
Madrid predicted SARS-CoV-2 positive deaths shall SARS-CoV-2 wasn’t deadly at all (i.e. IFR = 0.00)

Graphic A3.1.2 shows predicted SARS-CoV-2 positive deaths in Madrid shall the virus wasn’t deadly at all (i.e. IFR = 0). Herd immunity is reached regardless of the infection fatality rate. (Table A3.1 for all locations)

**Table A3.1.**
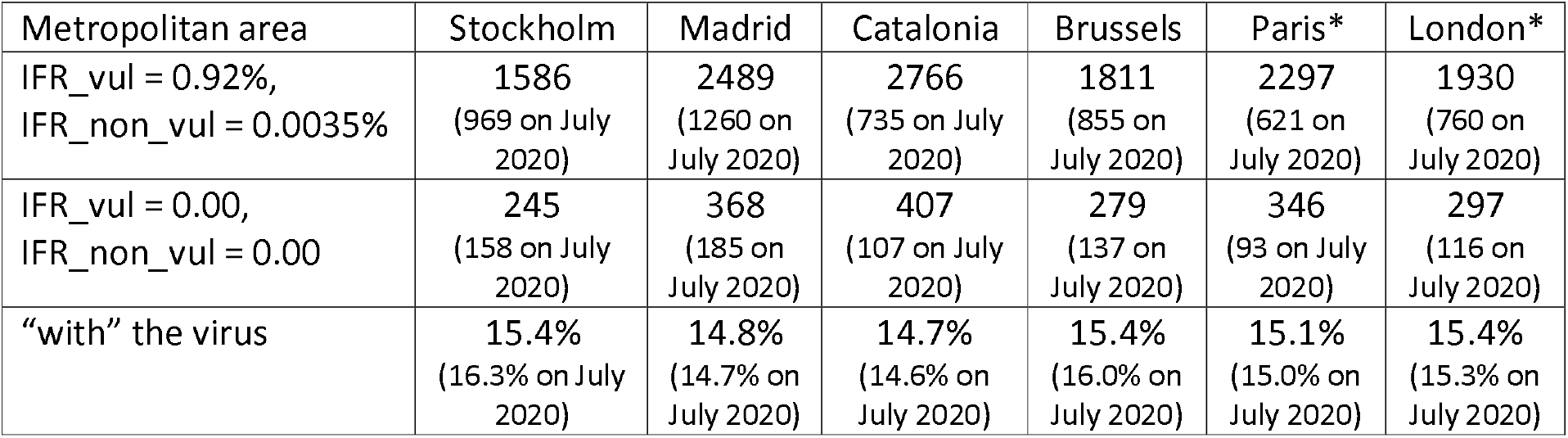
Final SARS-CoV-2 positive deaths per million (fitted isolations + normal life)

Since most deaths come from vulnerable individuals, proportion of reported SARS-CoV-2 positive deaths that were not caused by covid-19 is mostly invariant to isolation strategies (i.e. ≈ 15%). In a similar manner including SARS-CoV-2 tests, sensitivity and specificity would also lead to changes in the predicted SARS-CoV-2 positive deaths.

#### 2 days Do (infectiousness)

While fitting curves we came across that an exponentially decay 2 days infectiousness period (Do) coupled with a 5 days exposure (Eo) were able to explain the multiple valleys encountered after lockdowns in 1-day moving average daily deaths curves (fitting was done using 7-ma). We postulate that these valleys are not data reporting problems but inherent to the epidemic behavior (i.e. a feedback system with delay).

**Graphic A3.2.1.**
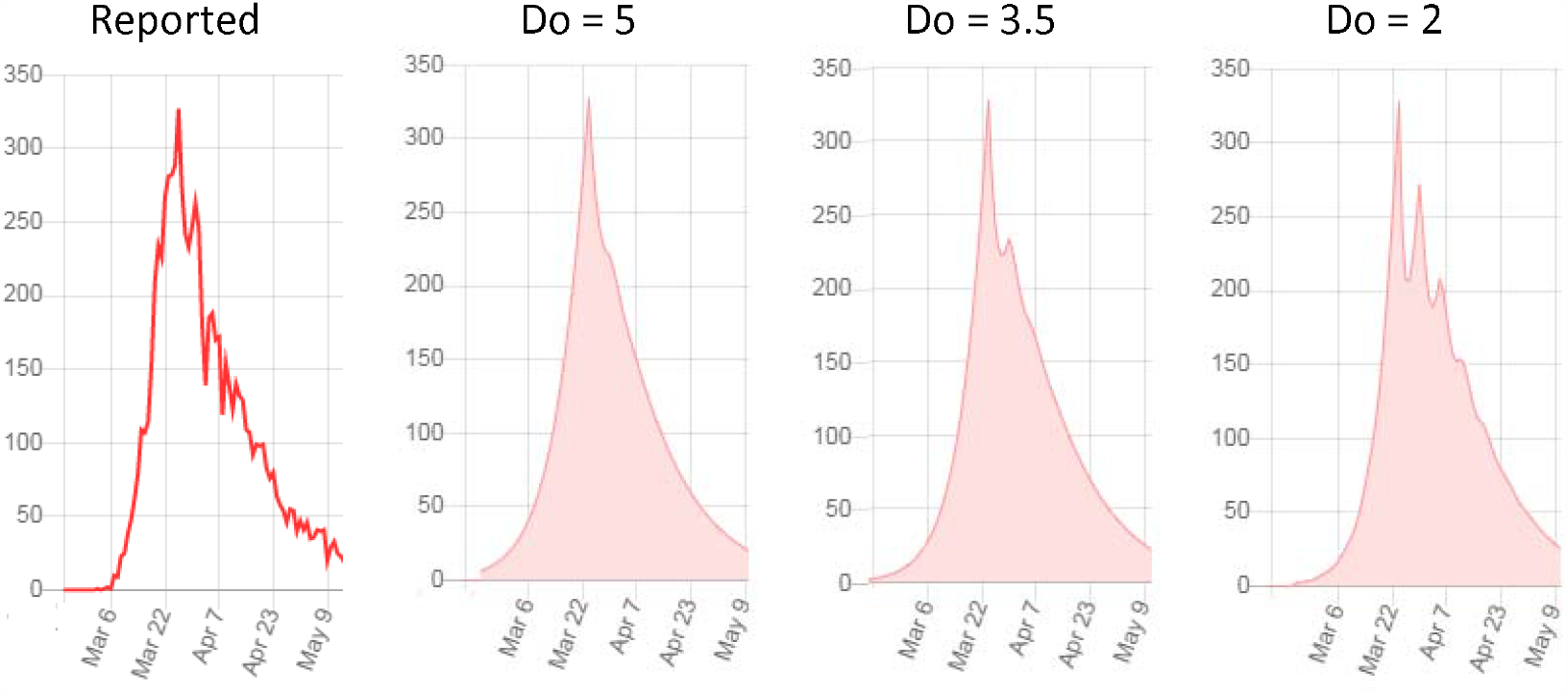
Madrid 1-day moving average predicted deaths for different Do infectiousness (Exposure Eo = 5)

**Graphic A3.2.2.**
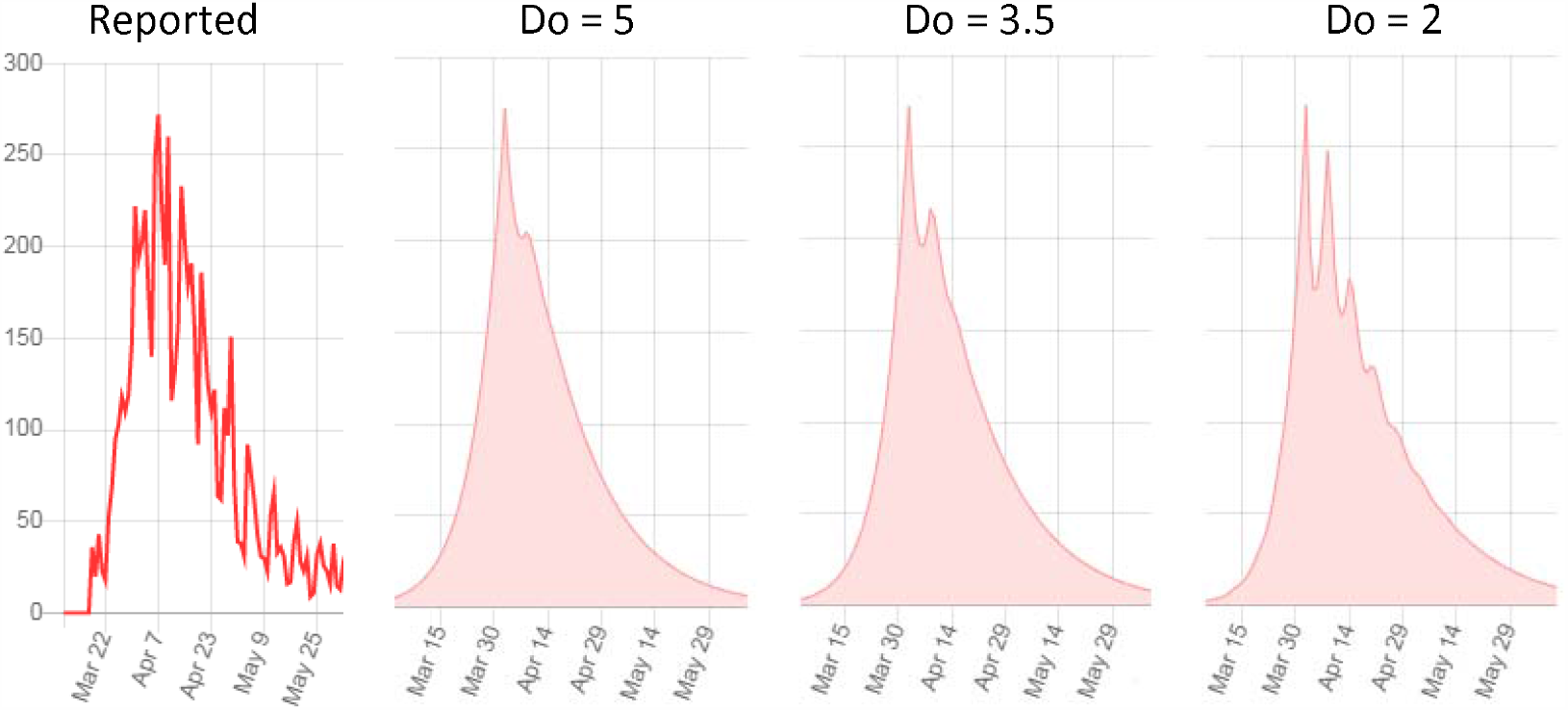
Paris 1-day moving average predicted deaths for different Do infectiousness (Exposure Eo = 5)

In (24) was found that the median period from infection to symptoms onset was 5.5 days (with fewer than 2.5% within 2.2 days and 97.5% within 11.5 days). In (25) was found that the serial interval (from exposure to infection of the secondary case) was 4 to 5 days and similar attack rates were found for symptomatic (1%) and presymptomatic (0.7%). In (26), where earliest swabs were taken on day 1 of symptoms, the authors stated that: “Critically, the majority of patients in the present study seemed to be beyond their shedding peak in samples from the upper respiratory tract when they were first tested”.

The simple formulae used here (i.e. nI[d] = nE[d-Eo] and nR[d] = I[d-1] / Do) imply Eo=5 days of zero contagiousness and an exponentially decaying infectiousness with average Do=2 days starting on day 6 (with 50% of contagious occurring within 6 days from exposure and 96.88% within 11 days). Real infectiousness function is likely to be more complex (e.g. not peaking on the first day of infectiousness), but we understand that Eo=5 and Do=2 are compatible with virus dynamic found in the field.

In (9) authors used Eo=5.1 and citing (26) set Do=5 (days an individual is assumed to be infectious on average). We understand findings in (26) establish that infections occurred during less than 5 days (peaking before) and not during 5 days on average. Under long enough isolation strategies, Do choice barely changes final immunity level (i.e. Ro and isolations levels determine it) but since Do does determine epidemic speed (i.e. it is the time scale) with short enough strategies lower values of Do will result in lower final immunity levels because HIT is reached during the isolation strategy and not once it has ended.

A hypothetical super spreading event (e.g. 1000 infected) not much before the beginning of lockdowns can send daily deaths into deeper valleys. If reported daily deaths are recorded exactly with the date of deceased, Paris and Stockholm 1-day moving average high variance data would be compatible with this close to lockdown dominant super spreading event. We maintain that for the formulae used here Do is lower than 3 because above that value, not even those close to lockdown super spreading events would be able to produce the observed multiple regularly spaced valleys.

We did not dig further to discard that daily patterns are just a weekend reporting problem, but it may be worth looking at it because such a short Do might also have implications for medical treatments.

#### Early D614 like strain wave in Asia: A hypothesis

Authors in (36) found that over the course of 1 month the new variant with Spike G614 reported in (37) replaced the original D614 as the dominant pandemic form. Also in (36), and even though G614 is associated with lower RT PCR Cts (suggestive of higher viral loads in patients), they did not find increased disease severity as measured by hospitalization outcomes.

Assuming that SARS-CoV-2 variant fitted here was the G614 (i.e. IFR_vul = 0.92%; Ro=3.3; Do = 2) and that the lower infectivity of the replaced variant D614 cannot only be the result of much lower Ro but also higher Do is needed (e.g. Ro_D614 = 2 and Do_D614 = 6), and also assuming the D614 variant does actually have much lower disease severity than G614 (i.e. IFR_vul_D614 = IFR_vul/5 = 0.18%); we modeled the two competing strains in Madrid with the G614 variant dominant spreading event on Dec 19 2019 (i.e. same date that fitted) and two different dates for the D614 dominant spreading events. (See in Appendix I: 2nd competing strain with complete cross-immunity).

Since we are hypothesizing lower IFR_vul for D614 in disagreement with (36) we refer to it as D614*

Results for the D614* arriving in Madrid on Aug 24 2019 are shown in Graphics A3.3.1. Even though it’s dominant spreader event occurs 117 days before G614’s, by March 2020 G614 dominates completely (D614* deaths are the small light blue curve). But if D614* arrives 60 days earlier (i.e. Jun 25, 2019) it manages to reach HIT before G614 has time to build its own wave. Since both competing strains are obtaining exposed individuals from the same susceptible compartment (i.e. complete cross-immunity) by the time G614 arrives it does not find enough susceptible to spread and final deaths are lower.

**Graphic A3.3.1.**
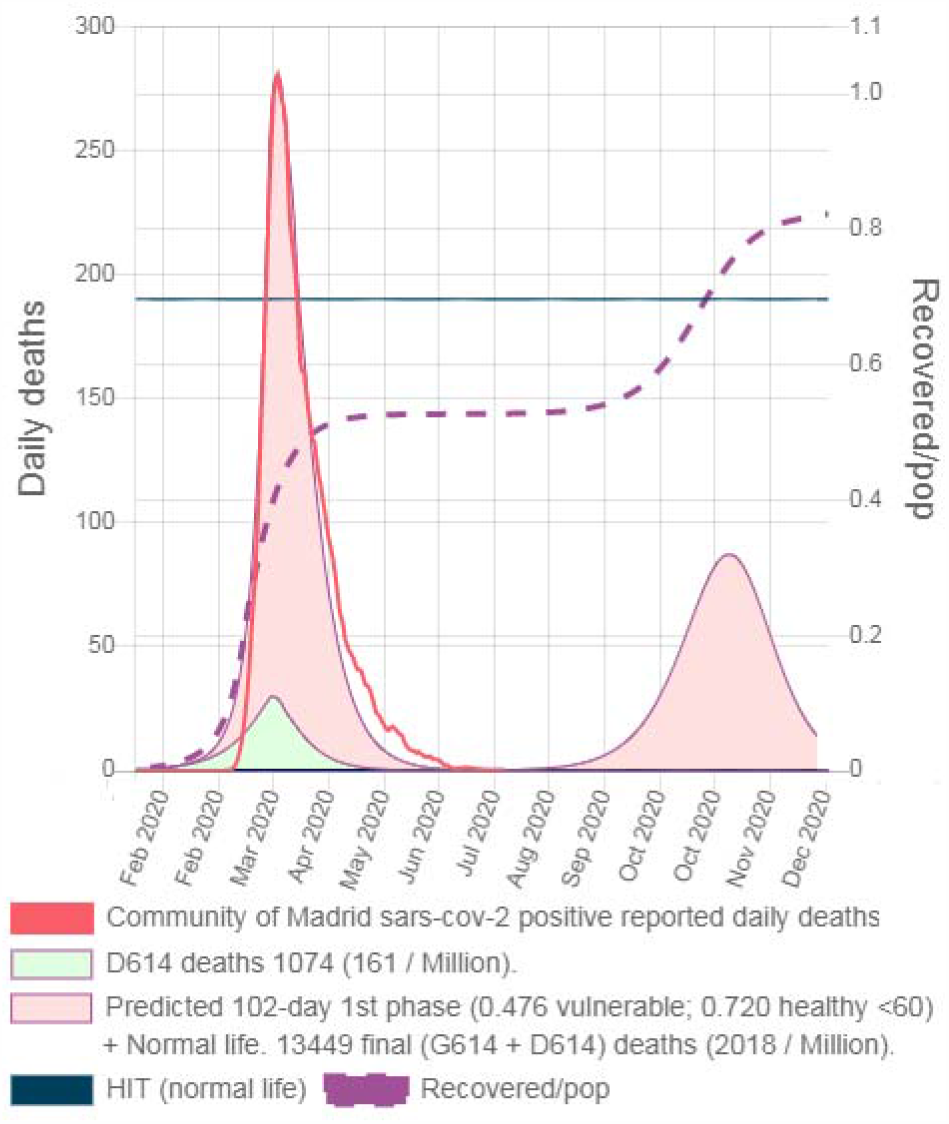
Madrid predicted SARS-CoV-2 positive deaths for SARS-CoV-2 variants G614 (Dec 19, 2019) and D614* (Aug 24 2019)

**Graphic A3.3.2.**
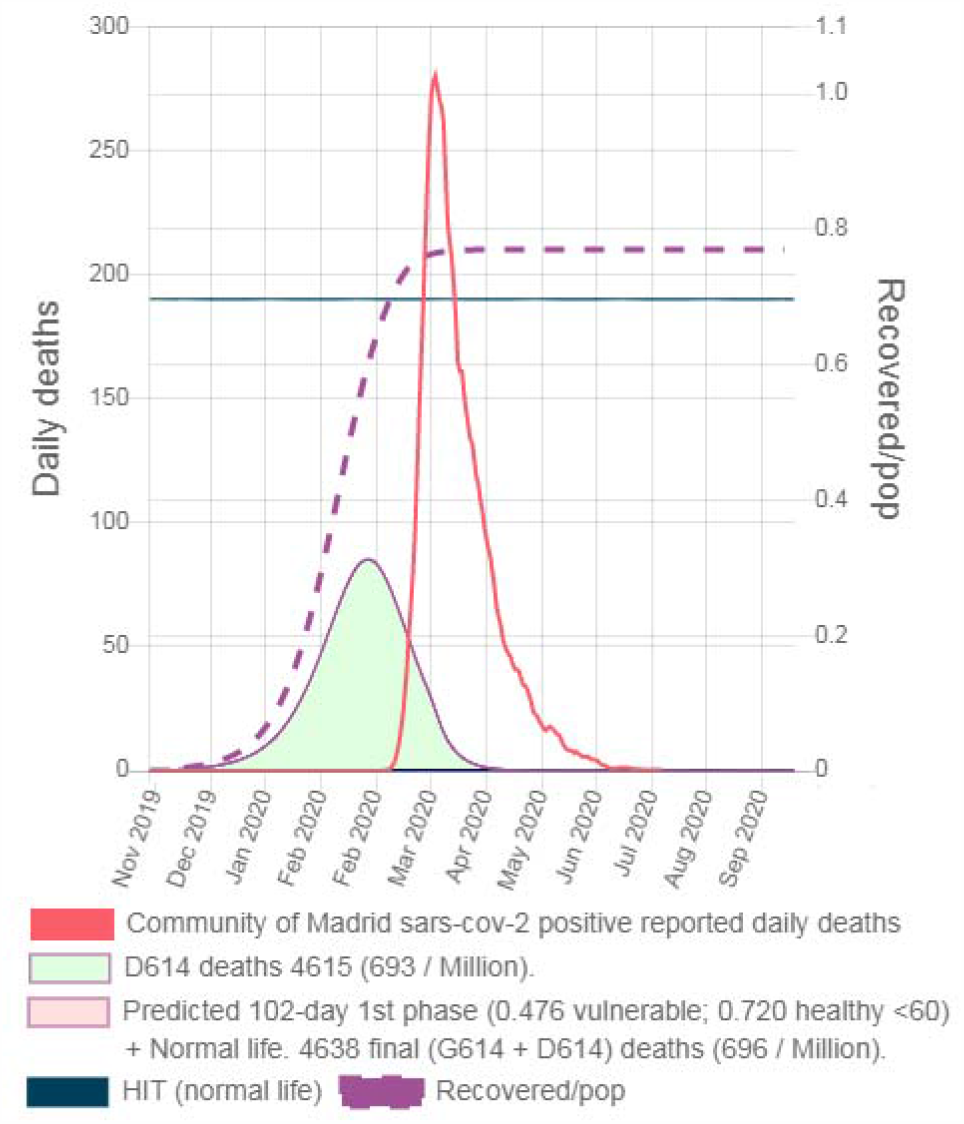
Madrid predicted SARS-CoV-2 positive deaths for SARS-CoV-2 variants G614 (Dec 19, 2019) and D614* (Jun 25 2019)

Note that since D614* has low infectivity it needs 5 months (from Jun 25 2019 to Nov 25 2019) to reach 11,201 infected in Madrid (i.e. 0.17% of the population). Thus for D614* the probability of one traveler from that location becoming a dominant spreader in another location remains low for many months. As neighbor locations are more connected, the probability of an early wave in Asian locations must have been higher than in Europe or America. Nevertheless, the probability of an Asian location of not having an early D614* wave would not be zero and analogously for a western location of do having it.

Thus the hypothesis that an early wave of a D614* like variant is the cause of the very low death count observed in Asian locations would be compatible with a non-negligible probability of finding some Asian locations in which death count was not very low (e.g. Wuhan) and western locations in which it was (e.g. Florianopolis, Brazil (41)).

The estimated Ro/Do ratio for G614 is 1.65 (i.e. 3.3/2) and that ratio must be much lower for the real D614 variant (e.g. 0.333) to G614 being able to rapidly overcome it. We set Do_D614 = 6 because trying to hypothesize a lower value (e.g. 4) would require Ro_D614 to be 1.333 to maintain the D614 ratio. With this low Ro, moving the D614* dominant spreading 7 months back would allow it’s wave to occur before the G614 arrival, but the immunity level achieved by the D614* wave would be lower (e.g. 45%) and thus the arrival of G614 (without isolations in place or without perfect timing with them) would result in a noticeable wave of deaths, and that did not occur.

#### Negligible reduction of HIT due to heterogeneity

Authors in (12) proposed that heterogeneity reduces HIT below 1-1/Ro level. We tested our 2-stratum model response for two plausible sources of heterogeneity: non-contagious asymptomatic reported in (42) and lower connectivity in young population as might be hinted in (8).

We modeled heterogeneity due to non-contagious asymptomatic^2^ (A) modifying the new exposed formulae with:

~~~
nE_abl[d] = S_abl[d-1]/pop * Ro/Do * (I_abl[d-1] * <u>((1-A_abl) + A_abl * A_inf)</u> * (1 - Iso_abl[d]) +
     + I_una[d-1] * <u>((1-A_una) + A_una * A_inf)</u> * (1 - auci));
nE_una[d]=S_una[d-1]/pop * Ro/Do * (I_una[d-1] * ((1-A_una) + A_una * A_inf) * (1 - Iso_una[d]) +
    + I_abl[d-1] * <u>((1-A_abl) + A_abl * A_inf)</u> * (1 - auci));
~~~

where A_inf: contagious asymptomatic (e.g. 0), A_abl: proportion of healthy <60 that are asymptomatic (e.g. 0.52 in Madrid), and A_una: proportion of vulnerable that are asymptomatic (e.g. 0.282 in Madrid). And heterogeneity due to lower connectivity^3^ (C) by increasing isolation to healthy <60 with the formula:

~~~
<u>level[j] = level[j] + (1-level[j]) * Iso_min;</u>
~~~

where level[j]: isolation level in day j; Iso_min: minimum isolation in normal life (e.g. Iso_min_abl = 0.4; Iso_min_una = 0.0).

If Ro=3.3 is used to fit reported daily deaths curves under the heterogeneous models, required infected on spread day drastically increase (e.g. in Madrid I_dayS = 1477.24 up from 1.10) and predicted deaths before lockdown depart too much from reported. See Graphics A3.4.1, A3.4.2 and A3.4.3.

**Graphic A3.4.1.**
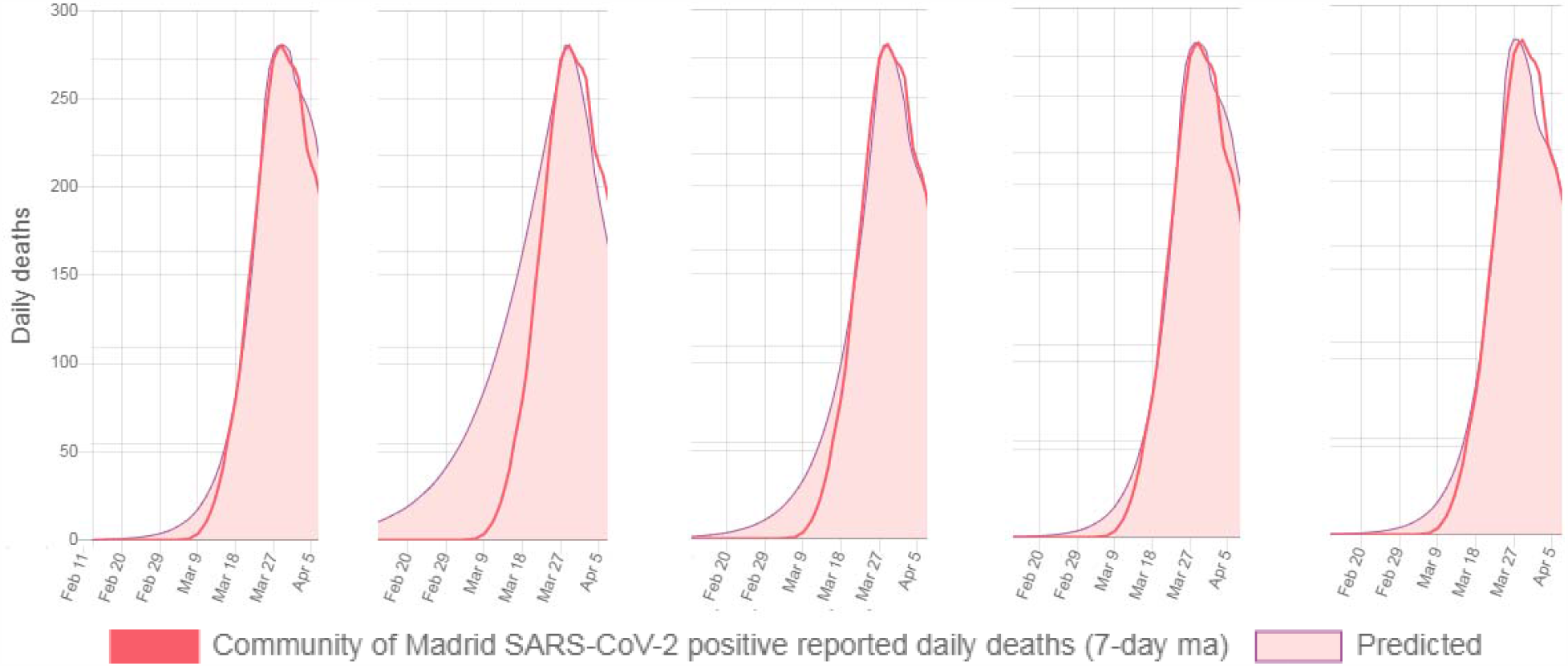
Homogeneity Ro = 3.3 I_dayS = 1.10 A3.4.2 Heterogeneity (A) Ro = 3.3 (RtoD=14) I_dayS = 1477.24 A3.4.3 Heterogeneity (C) Ro = 3.3 (RtoD=13) I_dayS = 40.24 A3.4.4 Heterogeneity (A) Ro = 5.912 I_dayS = 1.10 A3.4.5 Heterogeneity (C) Ro = 4.47 I_dayS = 1.10

This increase in I_dayS also occurs in every location (see Table A3.2). To maintain I_dayS = 1.10 in Madrid Ro must be increased to 5.912 for heterogeneity (A) and to 4.47 for heterogeneity (C). Fitting reported daily deaths curves and serology ratios using those Ro values for all locations also results in almost identical I_dayS values than under homogeneity with Ro=3.3 (see Table A3.2).

HIT levels were found searching uniform isolation values for vulnerable and healthy <60 for a 300-day long strategy starting on beginning of phase 1 that minimized final immunity level for a 7 years long simulation (see Table A3.2).

**Table A3.2.**
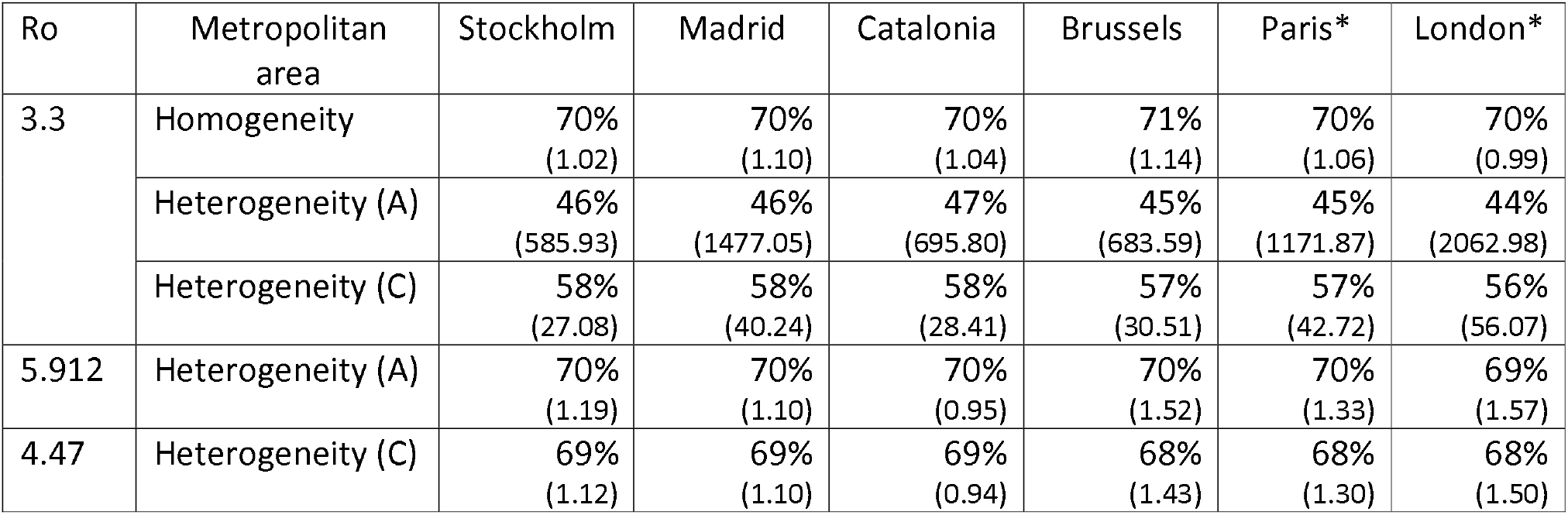
HIT under homogeneity and heterogeneity assumptions for different Ro and (infected on original spread day) necessary to fit reported daily deaths curves

| $R_0$ | Metropolitan area | Stockholm | Madrid | Catalonia | Brussels | Paris* | London* |
| --- | --- | --- | --- | --- | --- | --- | --- |
| 3.3 | Homogeneity | 70%<br>(1.02) | 70%<br>(1.10) | 70%<br>(1.04) | 71%<br>(1.14) | 70%<br>(1.06) | 70%<br>(0.99) |
|  | Heterogeneity (A) | 46%<br>(585.93) | 46%<br>(1477.05) | 47%<br>(695.80) | 45%<br>(683.59) | 45%<br>(1171.87) | 44%<br>(2062.98) |
|  | Heterogeneity (C) | 58%<br>(27.08) | 58%<br>(40.24) | 58%<br>(28.41) | 57%<br>(30.51) | 57%<br>(42.72) | 56%<br>(56.07) |
| 5.912 | Heterogeneity (A) | 70%<br>(1.19) | 70%<br>(1.10) | 70%<br>(0.95) | 70%<br>(1.52) | 70%<br>(1.33) | 69%<br>(1.57) |
| 4.47 | Heterogeneity (C) | 69%<br>(1.12) | 69%<br>(1.10) | 69%<br>(0.94) | 68%<br>(1.43) | 68%<br>(1.30) | 68%<br>(1.50) |

Shockingly, if Ro used under heterogeneity is increased (i.e. from 3.3 to 5.912 and 4.47) to properly fit reported deaths before lockdown, then HIT suffers almost no reduction from 70%.

Besides the caveats of using countrywide confirmed cases, authors in (12) fitted curves in Italy and Austria with CV=1 (i.e. coefficient of variation) using Ro=3, but somehow they managed to fit a high increase of heterogeneity (i.e. to CV=3) using almost the same Ro (i.e. maximum increase was 11%), and that might be the reason why they end up observing HIT reduction.

Homogeneity is implicitly assumed when Ro is estimated by fitting data at the beginning of the epidemic (e.g. Ro = 3.3). The increase of Ro (e.g. to 4.47) required to fit the same data when heterogeneity is assumed mostly compensates the reduction in HIT due to heterogeneity (see Table A3.2) even with substantial heterogeneity (i.e. Iso_min_abl = 0.4 >> Iso_min_una = 0.0). Therefore, we conclude that for SARS-CoV-2 reduction of HIT=1-1/Ro due to the plausible modeled heterogeneities is negligible.

Results must not be interpreted as heterogeneity not reducing HIT, because it might. For example HIT achieved under heterogeneity due to connectivity would be 59% if a high (maybe implausible) level of isolation to healthy <60 in normal life is assumed (e.g. Iso_min_abl = 0.8). In this case the required increase of Ro from 3.3 to 7.0 to fit the data does not completely compensate HIT reduction.

We also tested immunity level estimation and death minimizing isolation values under heterogeneity (see Tables A3.3 and A3.4). Immunity level estimation remained constant and minimum final deaths increased. Maximum values of isolation to vulnerable available for death minimizing under heterogeneity are the obtained while fitting Stockholm: 0.934 under heterogeneity (A) and 0.959 under heterogeneity (C).

“Back in March” death minimizing under both heterogeneity due to lower connectivity and due to non-contagious asymptomatic led to even lower values of isolations to healthy <60 than under homogeneity.

**Table A3.3.**
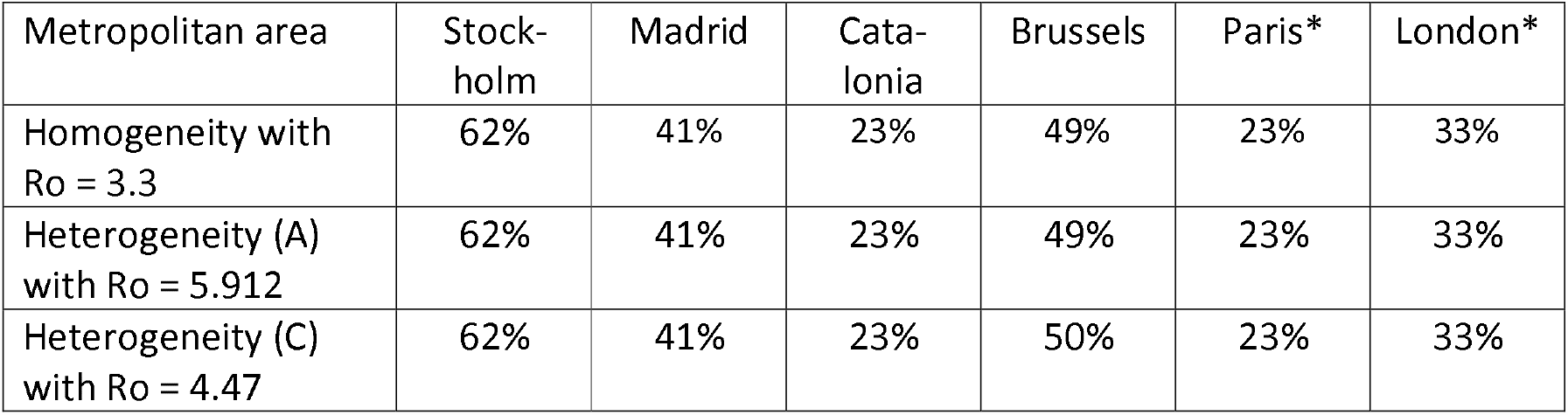
Immunity level estimation after fitted strategy under homogeneity and heterogeneity assumptions

**Table A3.4.** Final deaths per million for Back in March 2020 N-day death minimizing + NL under homogeneity and heterogeneity assumptions

| Metropolitan area | Stock-holm | Madrid | Cata-lonia | Brussels | Paris* | London* |
| --- | --- | --- | --- | --- | --- | --- |
| Homogeneity with $R_0 = 3.3$ | 1115<br>97-day<br>(0.941, +0.17) | 1269<br>102-day<br>(0.941, +0.16) | 1257<br>100-day<br>(0.941, +0.07) | 1011<br>101-day<br>(0.941, +0.23) | 1019<br>113-day<br>(0.941, +0.18) | 923<br>104-day<br>(0.941, +0.26) |
| Heterogeneity (A) with $R_0 = 5.912$ | 1186<br>97-day<br>(0.934, -0.05) | 1352<br>102-day<br>(0.934, -0.07) | 1370<br>100-day<br>(0.934, -0.22) | 1039<br>101-day<br>(0.934, +0.07) | 1083<br>113-day<br>(0.934, -0.03) | 952<br>104-day<br>(0.934, +0.11) |
| Heterogeneity (C) with $R_0 = 4.47$ | 1201<br>97-day<br>(0.959, -0.18) | 1372<br>102-day<br>(0.959, -0.21) | 1386<br>100-day<br>(0.959, -0.39) | 1057<br>101-day<br>(0.959, -0.05) | 1092<br>113-day<br>(0.959, -0.16) | 970<br>104-day<br>(0.959, 0.00) |

## Competing interests

All authors have completed the ICMJE uniform disclosure form at www.icmje.org/coi_disclosure.pdf and declare: no support from any organization for the submitted work; no financial relationships with any organizations that might have an interest in the submitted work in the previous three years; no other relationships or activities that could appear to have influenced the submitted work; Levan Djaparidze reports that he is the copyright holder of the model published in www.sars2seir.com.

## Footnotes

1 https://www.sars2seir.com/paper-12-2020/asia/

2 https://www.sars2seir.com/paper-12-2020/a0/

3 https://www.sars2seir.com/paper-12-2020/hc/

## Notes

### Funding Statement

No funding or finantial contributions to declare.

### Summary of Updates

This version was revised to correct the proportion of SARS-CoV-2 positive deaths reported in Brussels (from 1.00 to 0.69). It has impact on results (i.e. Brussels immunity level estimation decreased from 70% to 49% and therefore a second wave is predicted). Also the 2-stratum model responses for two plausible sources of heterogeneity were included in Appendix III: Negligible reduction of HIT due to heterogeneity.

